# Interpretable machine learning coupled to spatial transcriptomics unveils mechanisms of macrophage-driven fibroblast activation in ischemic cardiomyopathy

**DOI:** 10.1101/2025.08.18.25333841

**Authors:** Niranjana Natarajan, Hanxi Xiao, Shagufta Haque, Mary D Cundiff, Mika Hara, Varsha Sriram, Jishnu Das, Partha Dutta

## Abstract

Myocardial infarction (MI) often leads to ischemic cardiomyopathy, which is characterized by extensive cardiac remodeling and pathological fibrosis accompanied by inflammatory cell accumulation. Although inflammatory responses elicited by cardiac macrophages are instrumental in post-MI cardiac remodeling, macrophage microniche-mediated fibroblast activation in MI are not understood. Analyses of the spatial transcriptomics data of the hearts of patients with ischemic cardiomyopathy and a history of MI using a novel workflow combining Significant Latent Factor Interaction Discovery (SLIDE), which is an interpretable machine learning approach recently developed by us, regulatory network inference, and in-silico perturbations unveiled unique context-specific cellular programs and corresponding transcription factors driving these programs (that would have been missed by traditional analyses) in macrophages, and resting and activated cardiac fibroblasts. More nuanced analyses to examine the microniches comprising these cells in failed hearts uncovered additional cellular programs reflective of altered paracrine signaling among these cells. Silencing of niche-specific key genes and TFs from these cellular programs in both mouse and human macrophages altered the expression of pro-fibrotic genes. Furthermore, the secretomes from these macrophages suppressed myofibroblast differentiation. Finally, macrophage-specific *in vivo* silencing of *Tvp23b*, *Tdrd6,* and *B3galnt1* and the transcription factors *Mafk* and *Maz,* which are differentially expressed in macrophage/activated fibroblast niches, using a novel lipidoid nanoparticle approach in mice with MI significantly improved cardiac function and suppressed fibrosis. Our study uncovers novel macrophage niche-mediated fibroblast activation mechanisms and provides a new generalizable framework, coupling interpretable machine learning, regulatory network inference, in-silico perturbations, and *in vitro* and *in vivo* testing.

## Introduction

Heart failure (HF) characterized by inability of the myocardium to pump enough blood is the end stage clinical manifestation of a multitude of cardiovascular diseases (CVD), which is the leading cause of mortality worldwide and in the United States and a considerable burden on the healthcare system [1]. Characterized by the expansion of the cardiac interstitium through the disproportionate accumulation of extracellular matrix (ECM) proteins [2–4], pathological cardiac fibrosis is a hallmark of HF including ischemic cardiomyopathy [3, 5]. After MI, myofibroblasts promote ECM remodeling and fibrotic scar formation to prevent rupture of the ventricular wall [2, 6, 7]. Excessive fibrosis leads to adverse pathological cardiac remodeling and loss of compliance, contractility, and cardiac function, resulting in HF [2, 3]. A myriad of pro-fibrotic factors and pro-inflammatory cytokines like TGFβ, TNFα, and growth factors and the renin-angiotensin-aldosterone system (RAAS) are involved in the differentiation of fibroblasts into myofibroblasts [8–12]. Upon differentiation, myofibroblasts acquire new phenotypic characteristics, express contractile proteins (α-smooth muscle actin), and synthesize ECM proteins, matrix metalloproteinases (MMPs), and tissue inhibitors of MMPs (TIMPs) to remodel cardiac interstitium [9, 11].

Macrophages are the major resident immune cell population in the heart [13] and play a crucial role in adverse fibrotic remodeling through their paracrine effects on myofibroblasts [14, 15]. An ischemic injury to the heart stimulates an acute inflammatory response leading to recruitment of leukocyte populations from the bone marrow and spleen [16–18]. Pro-inflammatory monocytes of hematopoietic origin infiltrate the infarct area and mediate cardiac remodeling [14, 16, 18]. Elevated inflammatory biomarkers and increased macrophage infiltration in the LV have been associated with poor outcomes in HF patients [19–21]. However, clinical trials targeting inflammation in heart failure have been largely unsuccessful [22, 23].

While lineage tracing studies have identified the source and dynamics of cardiac fibroblasts in the injured heart [24, 25], the complexity of cardiac fibroblast subsets and the dynamic nature of fibroblast states present a major challenge in understanding molecular mechanisms underlying cardiac fibrosis after myocardial infarction (MI) [26–28]. Moreover, paracrine signaling between macrophages and fibroblasts shapes cardiac remodeling after MI as both cell types undergo sweeping changes in abundance, phenotype, and composition after an ischemic insult [11, 29, 30]. As such, understanding spatial relationships between these crucial cell types in the heart is pivotal to delineate the signals driving pathogenic fibroblast proliferation. Although recent studies employing single-cell omics techniques have shed light on transcriptomic and epigenetic states of individual cell types in the heart after MI, information on spatial niches, especially pathogenic functional microniches, is lost in these approaches. Spatial transcriptomic methods allow us to identify microniches and corresponding microniche-specific cellular programs that are directly linked to pathogenesis [31, 32].

Here, we employed GeoMx® whole transcriptome atlas to examine transcriptomic profiles of macrophages, resting fibroblasts, and activated fibroblasts in left ventricle specimens from patients with heart failure and history of MI and matched controls. We then developed a novel computational framework for the analyses of this spatial transcriptomic data coupling SLIDE, a novel interpretable machine learning approach that we recently developed [33], to regulatory network inference and in-silico perturbations. Using this framework, we identified cellular programs that were altered in different cell types (macrophages and resting and activated fibroblasts) as well as within cell types based on their spatial microniches (defined based on proximity to other cell types). Critically, several novel hits uncovered by our framework would have been missed by conventional analyses such as differential gene expression analyses. With a series of rigorous *in vitro* (murine and human primary macrophages and cardiac fibroblasts) and *in vivo* (a murine model of MI) experiments, we elucidated and validated the functions of key genes and TFs in the uncovered cellular programs. The key novel hits included *TVP23B*, *TDRD6,* and *B3GALNT1, MAFK,* and *MAZ*. These were expressed at high levels by macrophages closer to myofibroblasts and could reprogram resting fibroblasts into activated fibroblasts. More importantly, these novel functional hits were made possible by our systematic, multi-pronged approach for discovery (using machine learning), prioritization (guided by the systematic features of the computational analyses), characterization (*in vitro*), and validation (*in vivo*). This approach is generalizable across contexts and can be broadly applied as a discovery engine for spatial transcriptomic datasets.

## Results

### Spatial transcriptomic map of cardiac macrophages and fibroblasts in heart failure

Using spatial transcriptomics, we profiled macrophages, resting fibroblasts, and activated fibroblasts in human left ventricle specimens from four healthy donors and four patients with ischemic cardiomyopathy who had MI and underwent heart transplantation (**Figure 1A, S1A**). The latter group is referred to as HF hereafter. We captured 8-10 regions for sequencing per specimen and analyzed the transcriptome encompassing CD68+ macrophage regions, vimentin^+^ aSMA^−^ resting fibroblast regions and vimentin^+^ aSMA^+^ active fibroblast regions (**Figure 1A**: experimental design, **1B**: representative images). We sought to identify cellular programs, regulatory networks, and cellular communication altered in HF using a combination of novel interpretable machine learning approaches, regulatory network analyses, and in-silico perturbations. Specifically, we employed SLIDE, a novel machine learning approach recently developed by us, to identify unique transcriptomic signatures across cell types and spatial regions reflective of altered paracrine signaling in HF. We compared the transcriptomic signatures of (i) all cells, (ii) macrophages, and (iii) all, resting, and activated fibroblasts in heart failure vs. control, and (iv) macrophages proximal or distal to activated fibroblasts in heart failure. In each case, unique “latent factors” (context-specific co-expression modules) were identified using SLIDE. Critically, these programs would have been missed by conventional differential expression analyses, demonstrating the value of the interpretable machine learning approach. We then employed *in silico* perturbation analyses using Cell Oracle to identify key transcription factors in all cell types profiled in our analyses. Our SLIDE and Cell Oracle analyses prioritized a handful of key genes and transcription factors from the LFs (**Figure 1A**). These were assessed in mouse and human primary macrophages and cardiac fibroblasts using *in vitro* experiments. These experiments revealed the top 5 candidates (3 genes and 2 TFs), which were extensively characterized in a mouse model of MI-induced HF by silencing them specifically in macrophages using a novel lipidoid nanoparticles (**Figure 1A**).

**Figure 1:**
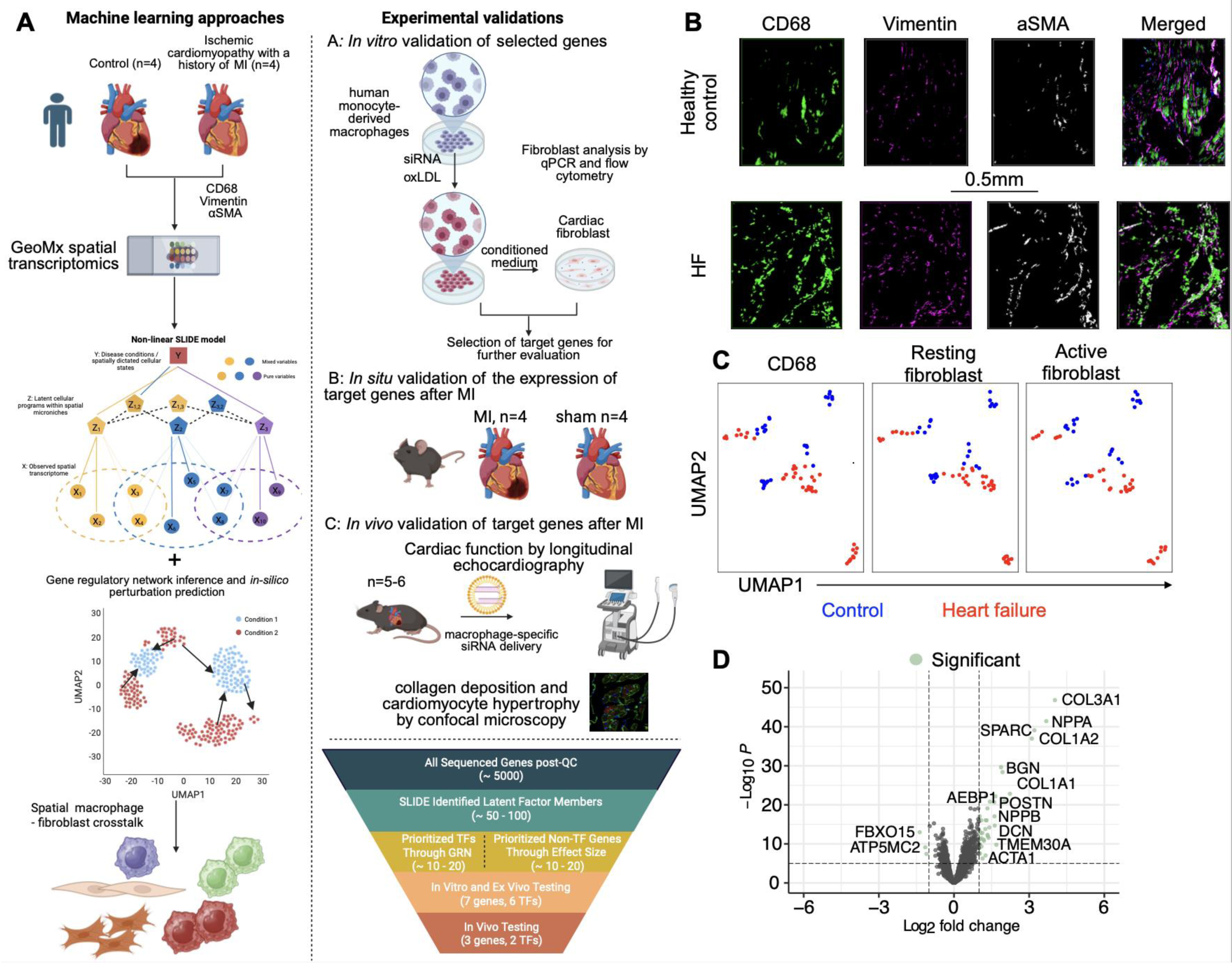
Interpretable machine learning approaches to uncover unique spatial microniches and gene regulatory networks of cardiac macrophages and fibroblasts in ischemic cardiomyopathy. Overview of the study (A), GeoMx digital spatial profiling (DSP) platform was used to examine the transcriptomic signature of cardiac macrophages, resting fibroblasts, and activated fibroblasts in ischemic heart failure samples and healthy controls. SLIDE, a novel machine learning approach, was employed to identify unique latent factor modules in heart failure, and gene regulatory networks were analyzed with Cell Oracle. Genes and transcription factors of interest identified by the computational analyses were validated (a) *in vitro* in human monocyte-derived macrophages and cardiac fibroblasts, (b) *in situ* using sham and MI hearts, and (c*) in vivo* by selective gene silencing in macrophages in mice with MI. B: Representative images (immunostaining) of the capture areas of GeoMx DSP are shown. C: UMAP visualization shows distinct clusters of heart failure and control macrophages (CD68), resting fibroblasts, and activated fibroblasts. D: Volcano plot of differentially expressed genes in all cells in heart failure vs. control. n=4/group.

### Global transcriptomic differences across HF and control

We first focused on elucidating global differences across HF and control. After processing through a standard analytic pipeline including rigorous quality control (Methods), we retained ∼4,000 genes for downstream analyses (**Figure S1B**). Unsupervised analyses of the spatial transcriptomes (**Figure 1B**) revealed differences across cell types between HF and control (**Figure 1C**). As anticipated, conventional differential expression (DE) analyses comparing transcriptomic signatures of all cell types revealed several fibrotic and extracellular matrix (ECM) genes to be upregulated in HF compared to healthy controls (**Figure 1D**). Periostin (*POSTN*), vimentin (*VIM*), skeletal alpha actin (*ACTA1*), fibronectin (*FN1*), SPARC, and the collagen isoforms 1A1, 1A2, 3A1, 6A1, 6A2, and 6A3 were enriched in HF (**Figure 1D**). However, these analyses are non-cell-type-specific. Hence, we next delved into elucidating cell-type-specific pathological cellular programs

### Macrophage-specific pathological cellular programs

To identify differences in gene expression signatures within the macrophage population between HF vs. control, we first used DE analyses. We identified a small number of genes with higher expression in HF macrophages **(Figure 2A**). As expected, a few profibrotic and ECM component genes (*COL1A1, COL1A2, COL3A1*, and *SPARC*) had significantly increased expression in HF relative to control **(Figure 2A**). We also observed higher expression of *NPPA*, which encodes atrial natriuretic peptide, in HF macrophages, consistent with findings of this gene being associated with higher risk of HF [34],[35, 36]. However, while interesting and in line with expectation, these analyses identified only a small number of DE genes as these univariate analyses are relatively underpowered.

**Figure 2:**
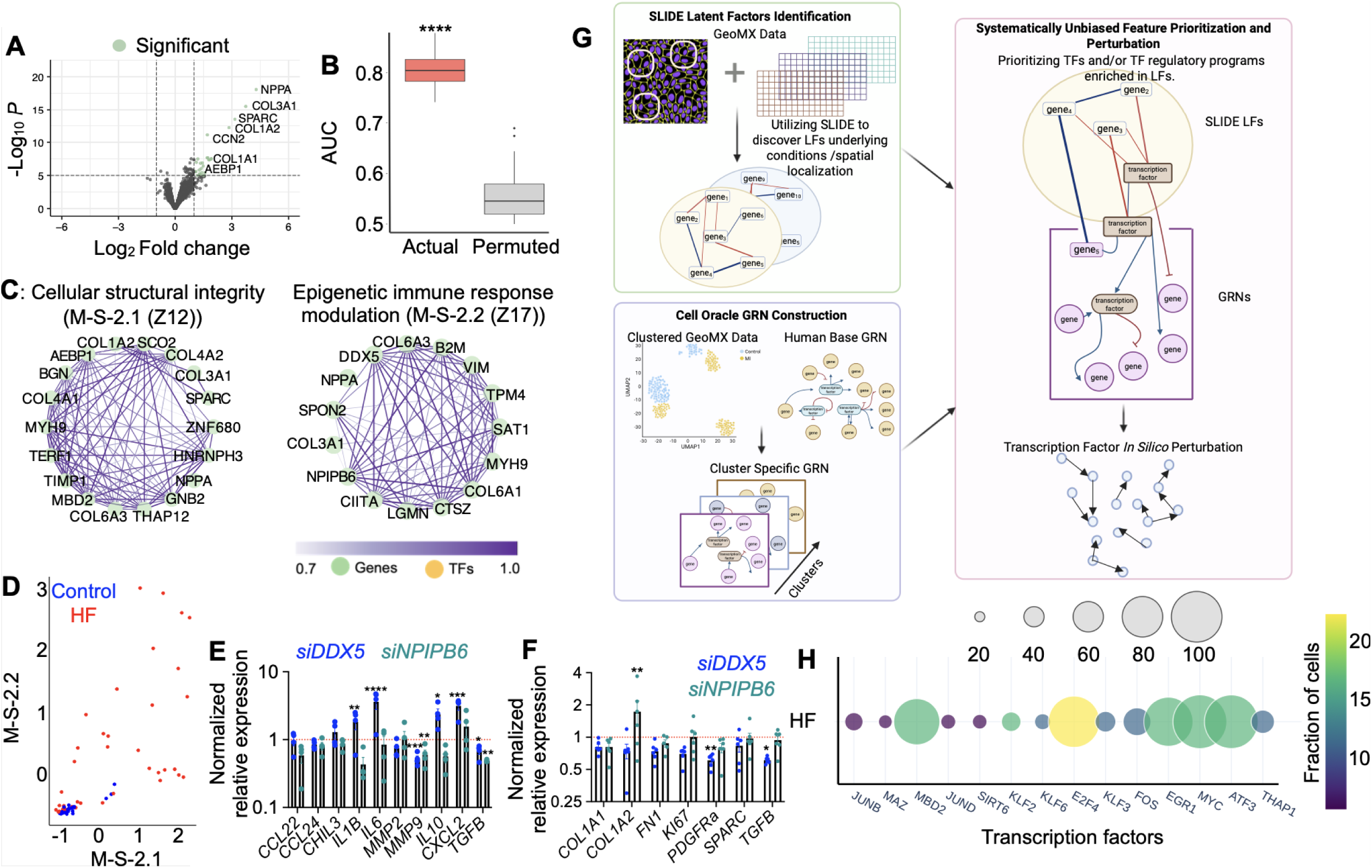
Identification of dysregulated macrophage-specific cellular programs in heart failure using SLIDE. A: Volcano plot of differentially expressed genes in HF macrophages compared to control cardiac macrophages. B: SLIDE model discriminates HF and control. C: Significant standalone LFs visualized as correlation networks-cellular structural integrity (M-S-2.1), and epigenetic immune response modulation (M-S-2.2). D: Scatter plot visualizing stratification of HF vs control cardiac macrophages in latent space (M-S-2.1 and M-S-2.2). E: Gene expression by human monocyte-derived macrophages after *DDX5* and *NPIPB6* silencing. F: The expression of the genes was determined by qPCR in human cardiac fibroblasts cultured with the media from human monocyte-derived macrophages treated with the siRNA. G: Gene regulatory networks in cardiac macrophages were identified using Cell Oracle using a combination of gene regulatory network construction and *in silico* perturbations (schematic). H: Key transcription factors controlling macrophage cellular programs in heart failure prioritized using regulon enrichment and in-silico perturbation analyses. *p<0.05, **p<0.01, ***p<0.001, ****p<0.0001. B: E,F: one-way ANOVA with post-hoc Fisher LSD test. n=4/group.

To gain more comprehensive insights into complex cellular programs altered in HF, we employed SLIDE, a novel interpretable machine learning approach we recently developed [33]. A key innovation of SLIDE is the identification of context-specific latent factors (in this case, modules of co-expressed genes) that are necessary and sufficient to infer differences across the groups being compared (here HF and control). SLIDE identified five latent factors that provided significant discrimination between HF and control cardiac macrophages. (**Figures 2B and S2A**). The latent factors are visualized as correlation networks (co-expression modules). Of the 5 latent factors, M-S-2.1 and M-S-2.2 are (for naming conventions of latent factors, please see Methods) significant standalone latent factors i.e., those that provided the maximum discrimination – the linear part of an overall non-linear model (**Figure 2C**). They encompassed cellular structural integrity/ECM organization as reflected by the enrichment of ECM genes such as *COL1A2, COL3A1, COL4A2, COL6A1, COL6A3* and key regulators of cellular structural integrity such as *MBD2* [37, 38] and *AEBP1* [39, 40] (**Figure 2C**). Significant standalone latent factors also comprised of the genes involved in epigenetic immune response modulation, as evidenced by the high expression of *CIITA*. Critically, SLIDE uncovered these well-characterized mechanisms without incorporating any prior knowledge, demonstrating the robustness and power of our interpretable machine learning approach in capturing biologically meaningful cellular programs. However, in addition to recovering known mechanisms, SLIDE also captured several novel candidates including *CIITA, DDX5*, and *NPIPB6* to be enriched in HF macrophages. Overall, the expression programs captured by just these two latent states provided excellent discrimination between HF and control (**Figure 2D**). In addition to these 2 standalone latent factors, SLIDE also identified 3 significant interacting latent factors i.e, the signatures that add to the significant standalone factors – the non-linear part of the model. These latent factors include M-I-E-2.1, M-I-E-2.2 and M-I-E-2.3, which reinforced the fibrotic signature and reflected altered immune signaling (**Figures S2B-D**).

We have demonstrated across contexts [41–44] that SLIDE can provide putative mechanistic insights beyond the inference of biomarkers alone. Thus, we sought to validate the function of both key known and novel genes identified by SLIDE analyses using an *in vitro* approach (**Figure 1A**). The prioritization of these genes was done using a set of systematic criteria based on contributions of the genes to the identified significant latent factors (loadings on the allocation matrix) and their individual effect sizes. Within the significant latent factors, we prioritized genes with high contributions and/or high individual effect sizes (Methods). This provides a systematic (without cherry picking) way to identify downstream hits for functional validation. We examined profibrotic gene expression after silencing the genes of interest in macrophages. Macrophage-fibroblast paracrine signaling was examined by treating cardiac fibroblasts with conditioned medium from macrophages after gene silencing. To validate the hits obtained from the SLIDE analysis, we silenced the genes, such as *Aepb1*, *Ciita,* and *Mbd2*, for which there is already evidence of association with cardiac remodeling. Macrophage *Mbd2* has been identified to promote pulmonary and renal fibrosis by upregulating TGFB production [37, 38]. *Aebp1* has been shown to promote hepatic, renal, and cardiac fibrosis [39, 45], and elevated circulating levels of AEBP1 correlated with higher cardiac fibrosis in HF patients [39]. We first tested these three genes in an orthogonal system (cultured murine bone marrow-derived macrophages, BMDMs) (**Figure S2E-G**). The expression of *Mmp2* and *Mmp14*, which are crucial in cardiac remodeling in HF [46–48], significantly diminished after the gene silencing. Interestingly, the expression of pro-fibrotic interleukins *Il10*, *Il15*, and *Il25*, was elevated in BMDM treated with si*Mbd2* (**Figure S2F**). However, murine cardiac fibroblasts treated with the conditioned medium from the BMDM did not exhibit significant changes in the expression of collagen isoforms and pro-fibrotic genes (**Figure S2G**).

Next, we focused on two novel genes identified in the latent factors –*DDX5* and *NPIPB6*. To characterize their roles, we silenced them in human monocyte-derived macrophages (HMDM). We observed alterations in pro-inflammatory and pro-fibrotic markers in the macrophages after si*DDX5* treatment (**Figures 2E and S2E**). Cardiac fibroblasts treated with the conditioned medium obtained from MDM treated with si*DDX5* exhibited reductions in *TGFB* and *PDGFRa* expression (**Figure 2F)**. Interestingly, the conditioned medium derived from si*NPIPB6*-treated MDM increased *COL1A2* expression in fibroblasts (**Figure 2F**). Together, these results demonstrate that SLIDE recapitulates key known and novel genes with functional significance.

To better dissect transcription factors (TFs) and corresponding regulatory circuits controlling these cellular programs identified by SLIDE, we adapted Cell Oracle to reconstruct cell-state-specific gene regulatory networks (GRNs) for these macrophages [49]. Briefly, we used base GRNs derived from Cell Oracle and fit context-specific GRNs using a Bayesian ridge regression approach on the spatial transcriptomic data (**Figure 2G**) to form cluster (cell-state)-specific GRNs. We then tested for enrichment of specific TF regulons, regulatory modules including genes downstream of TFs in the GRN, in the significant standalone and interacting latent factors identified by SLIDE, coupled with *in silico* perturbation (**Figure 2H**). A TF was prioritized only if it passed both criteria – a regulon downstream of the TF was enriched in the latent factor and *in silico* perturbation of the TF using Cell Oracle had a significant predicted effect relative to a random permutation. Intriguingly, although SLIDE does not use any prior knowledge, the latent factors identified by SLIDE are enriched for regulons downstream of several key transcription factors. Several of these TF regulons had significant inferred functional consequences using *in silico* perturbations. TFs prioritized using the two criteria mentioned above included MDB2, E2F4, EGR1, MYC, and ATF3 (**Figures 2H and S2H)**. This demonstrates that we capture important regulons in the macrophage-specific GRN that discriminate HF and controls. This also highlights the biological relevance of the non-linear model. While the linear component of the model (the significant standalone latent factors) provides excellent discrimination between the patient groups, the significant interacting latent factors help infer aspects of GRN architecture driving the cellular programs. Overall, our analyses provide insights into macrophage cellular programs and associated TF regulons that drive differences between HF and control.

### Cardiac fibroblast specific cellular programs in heart failure

Next, to understand altered fibroblast cellular programs in HF, we analyzed the transcriptomic signature of all fibroblasts, resting fibroblasts, and activated fibroblasts in HF. Using DE analyses, as expected, we observed an upregulation of various collagen isoforms, *POSTN*, *ACTA1*, and *AEBP1* (**Figure 3A**). However, as in macrophages, these analyses only identified a small number of DE genes as DE analyses are relatively underpowered. A SLIDE model however provided significant discrimination between HF and control fibroblasts (**Figure 3B**). The model included one standalone significant latent factor-F-S-3.1 (**Figure 3C**) and three significant interacting latent factors – F-I-E-3.1, F-I-E-3.2, and F-I-E-3.3 (**Figure S3A**). The significant standalone latent factor F-S-3.1 by itself provided excellent stratification between HF and control (**Figure 3D**) and reflected alterations in collagen synthesis/ECM genes (**Figure 3C**). The additional interacting latent factors cellular programs reflected changes in cardiac function, cellular metabolism and homeostasis, and ribosome biogenesis (*COL4A1*, *VIM*, and *B2M*), (**Figures S3B-D)**.

**Figure 3:**
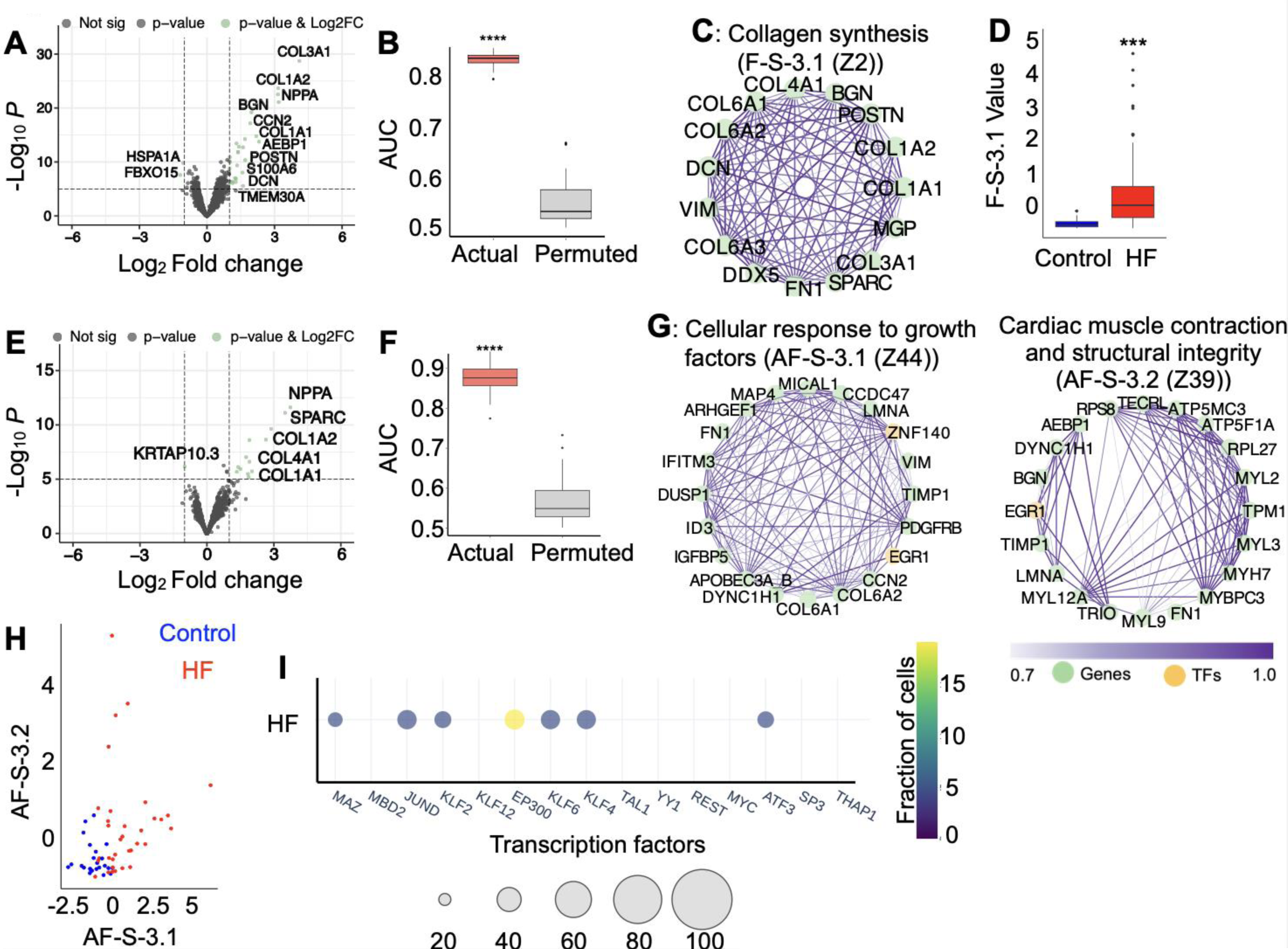
Dysregulated fibroblast-specific cellular programs in heart failure. A: Volcano plot of all fibroblast-specific DEGs in HF vs. control. B: SLIDE model discriminates fibroblast signatures in heart failure vs control. C: Significant standalone LF is visualized as a correlation network-collagen synthesis (F-S-3.1). D: SLIDE model is applied to resting fibroblasts in HF vs. control. The model performance is shown in the box plot. E: Volcano plot of differentially expressed genes in HF vs. control activated fibroblasts. F: SLIDE model performance in activated fibroblasts. G: Significant standalone latent factors in activated fibroblasts are visualized as correlation networks, cellular response to growth factors (AF-S-3.1) and cardiac muscle contraction and structural integrity (AF-S-3.2). H: Scatter plot visualizing stratification of HF vs. control activated fibroblasts in latent space (AF-S-3.1 and AF-S-3.2). I: Key transcription factors controlling activated-fibroblast-specific cellular programs in heart failure prioritized using regulon enrichment and in-silico perturbation analyses. ***p<0.001, ****p<0.0001

However, these signatures reflect transcriptional states encompassed by both resting and activated fibroblasts. To elucidate more specific molecular signatures of pathological cardiac fibrosis, we next examined the transcriptomic signatures of resting fibroblasts and activated fibroblasts separately. In resting fibroblasts, *COL1A1, COL1A2*, *COL3A1, FN1*, and *POSTN* were upregulated in HF based on DE analyses (**Figure S4A**). However, as earlier, DE analyses were underpowered at teasing apart differences. A SLIDE model focused only on resting fibroblasts was significantly discriminative between HF and control (**Figures S4B-C)**. However, the transcriptional signatures primarily included cellular processes crucial in fibrotic remodeling and associated immune changes (**Figures S4D-K**).

More interestingly, we saw more dramatic transcriptional differences in the activated fibroblasts. Like total fibroblasts, DE analyses in activated fibroblasts show that the collagen isoforms and SPARC were upregulated (**Figure 3E**). More interestingly, using SLIDE, we obtained a model that was significantly discriminatory between activated fibroblasts from HF and control individuals. We identified two key standalone significant latent factors (modules of co-expressed genes) and five interacting latent factors that distinguished between HF and controls (**Figures 3F-G and S5**). The two significant standalone latent factors, AF-S-3.1, AF-S-3.2, by themselves, provided excellent stratification between HF and control (**Figure 3H**). Interestingly, AF-S-3.1 and AF-S-3.2 reflect processes beyond ECM reorganization including cellular response to growth factors, cardiac muscle contraction, and structural integrity (**Figure 3G**). Genes in these significant LFs include AEBP1, PDGFRB, vimentin, fibronectin, ZNF140, mitochondrial proteins ATP5MC3 and ATP5F1A, collagen isoforms, and TIMP1. Mitochondrial mechanisms and metabolism play crucial roles in myofibroblast differentiation and fibrosis [26], ATP5MC3 and ATP5F1A are components of mitochondrial complex V. Finally, we sought to examine the underlying transcription factors driving these molecular signatures in activated fibroblasts in heart failure. To this end, we performed enrichment analysis for corresponding regulons, which includes the interacting latent factors, and in-silico perturbations as described earlier. We found that the transcription factors EP300, KLF4, and KLF2 drive regulatory programs in these fibroblasts (**Figure 3I**).

### Macrophage-fibroblast spatial microniches in heart failure

To further delve into microenvironment and paracrine signaling between macrophages and activated fibroblasts in heart failure, we examined differences in transcriptomic signatures of macrophages proximal (based on spatial density of nuclei – see Methods) to activated vs. resting fibroblasts in heart failure specimens. Only *HHATL* (Hedgehog Acyltransferase Like), which, mediates N-terminal palmitoylation of target proteins and is involved in hedgehog signaling and associated with hypertrophic cardiomyopathy [50], was significantly upregulated in macrophages proximal to activated fibroblasts (**Figure 4A**). As such, differential expression analyses did not yield significant insights into the alterations in transcriptomic state of the macrophages based on their locations, underscoring the limited power of univariate approaches in elucidating these more complex differences. However, a SLIDE model was able to significantly discriminate these regions in HF samples, demonstrating that it is powerful at identifying complex cellular programs in spatial microniches (**Figure 4B**). We identified one significant standalone and two significant interacting latent factors that distinguished cardiac macrophages proximal to activated fibroblasts compared to resting fibroblasts-MD-S-4.1, MD-I-E-6.1, MD-I-E-6.2 (**Figure 4C, Figure S6A-B**). The significant standalone latent factor, MD-S-4.1 by itself provided excellent stratification between the macrophages from these two microniches (**Figure 4D**). This complex cellular program reflects differences that are solely based on spatial microenvironment (proximity to activated fibroblasts vs. lack thereof) within the same condition (HF) and cell type (macrophages). We hypothesized that this novel program captures differences in spatial microniches and corresponding paracrine signaling.

**Figure 4:**
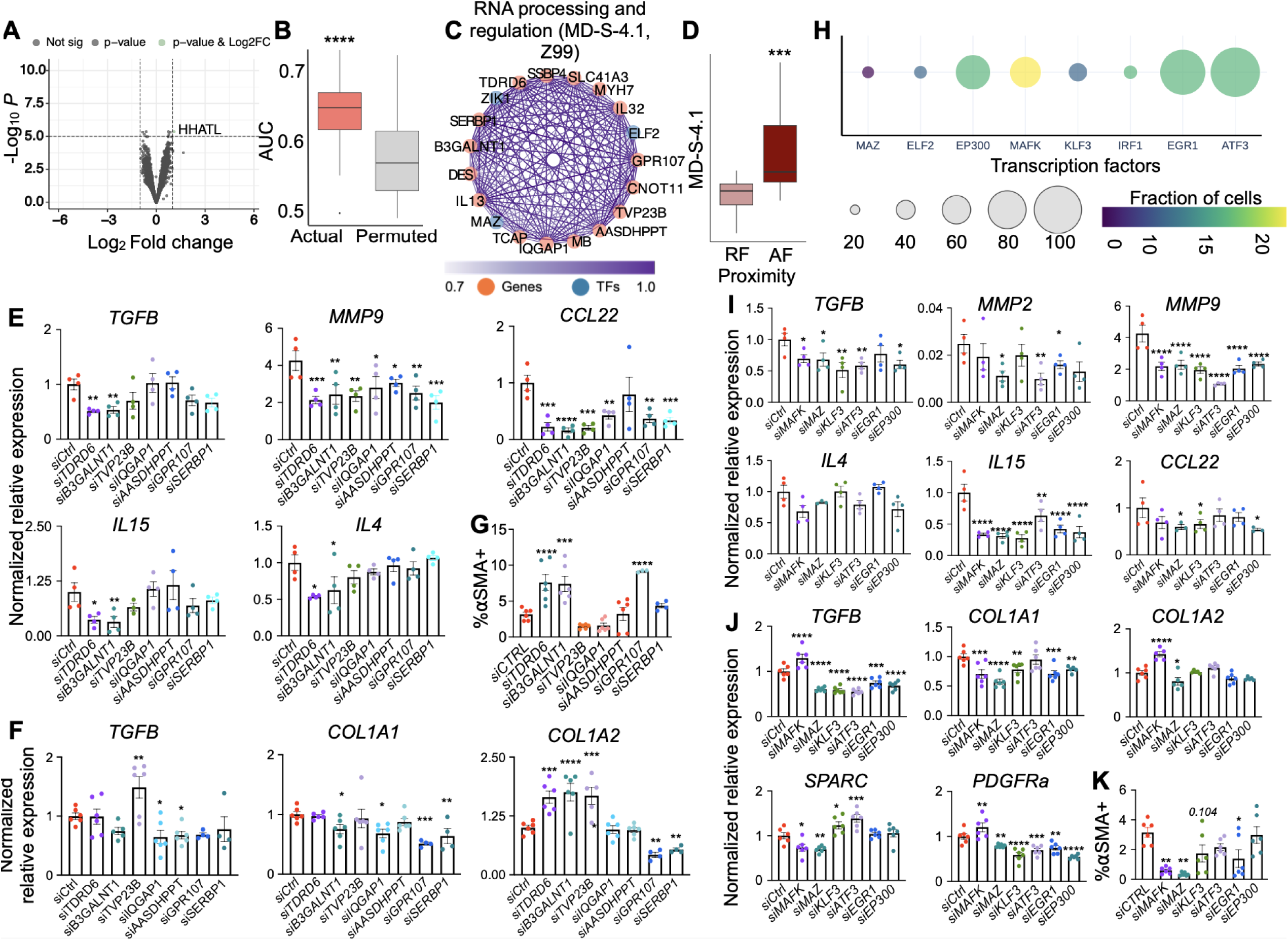
Identification of spatial microniches of macrophages in heart failure patients based on proximity to activated vs. resting fibroblasts. A: Volcano plot showing DEGs in macrophages proximal to activated vs. resting fibroblasts in HF patients. B: SLIDE model performance discriminating macrophages in HF specimens proximal to activated vs. resting fibroblasts. C: Significant standalone LF visualized as a correlation network, RNA processing and regulation (MD-S-4.1). D: Latent factor MD-S-4.1 distinguishes macrophages proximal to activated fibroblasts (AF) compared to the ones proximal to resting fibroblasts (RF). E: The expression of the pro-fibrotic genes was assessed in HMDM after silencing *TDRD6, B3GALNT1, TVP23B, IQGAP1, AASDHPPT, GPR107*, and *SERBP1*. F: The expression of the genes was determined by qPCR in human cardiac fibroblasts cultured with the media from human monocyte-derived macrophages treated with the siRNA. G: ⍺SMA^+^ fibroblasts were quantified by flow cytometry in human cardiac fibroblasts cultured with the media from human monocyte-derived macrophages treated with the siRNA. H: Key transcription factors controlling these spatial niche-specific cellular programs prioritized using regulon enrichment and *in silico* perturbation analyses. I: The expression of the genes was determined by qPCR in human monocyte-derived macrophages treated with the siRNA against the TFs. J: The expression of the genes was determined by qPCR in human cardiac fibroblasts cultured with the media from human monocyte-derived macrophages treated with the siRNA. K: ⍺SMA^+^ fibroblasts were quantified by flow cytometry in human cardiac fibroblasts cultured with the media from human monocyte-derived macrophages treated with the siRNA. *p<0.05, **p<0.01, ***p<0.001, ****p<0.0001. E, F, G, I, J, and K: one-way ANOVA with post-hoc Fisher LSD test. n=4-5/group.

From the list of genes in our significant latent factors, we further prioritized *TDRD6*, *B3GALNT1*, *TVP23B*, *IQGAP1*, *AASDHPPT*, *GPR107*, and *SERBP1* in human monocyte-derived macrophages and cardiac fibroblasts based on the same systematic criteria described above (contributions to these significant latent factors and individual effect sizes). The functions of most of these genes in the context of cardiovascular disease are not well known. These genes were silenced in human macrophages (**Figure S6C**). Pro-fibrotic gene expression (*TGFB, MMP9, CCL22, IL15,* and *IL4*) was significantly altered upon silencing several of the genes of interest (**Figure 4E**). In cardiac fibroblasts treated with the conditioned medium derived from the macrophages after the gene silencing, we observed increased expression of *TGFB* in si*TVP23B*, decreased *TGFB* levels in si*IQGAP1* and si*AASDHPPT*, and diminished *COL1A1* in si*B3GALNT1*, si*IQGAP1*, si*GPR107,* and si*SERBP1* treatments (**Figure 4F**). Interestingly, *COL1A2* levels were increased in si*TDRD6*, si*B3GALNT1,* and si*TVP23B* treatments while *COL1A2* expression was suppressed after *GPR107* and *SERBP1* silencing (**Figure 4F**). Finally, we examined aSMA expression in cardiac fibroblasts by flow cytometry. Conditioned medium from si*TDRD6*, si*B3GALNT1* and si*GPR107*-treated macrophages increased the frequency of ⍺SMA^+^ fibroblasts, compared to siControl (**Figure 4G**).

Lastly, we assessed the transcription factors that drive differences in regulatory architectures of cardiac macrophages proximal to activated vs. resting fibroblasts. GRN construction followed by regulon enrichment and *in silico* perturbation analyses using Cell Oracle indicate that the transcription factors MAZ, EP300, MAFK, KLF3, IRF1, EGR1, and ATF3 are crucial for macrophages to receive different paracrine signals based on their proximity to activated fibroblasts. (**Figures 4H and S6D**). We next silenced these TFs in HMDM and observed significant downregulation of *MAFK, MAZ, KLF3, ATF3,* and *EP300* (**Figure S6E**). Except *TGFB* in si*EGR1*, *TGFB, MMP9*, and *IL15* were significantly decreased in all conditions compared to siControl (**Figure 4I**). Cardiac fibroblasts treated with conditioned medium from a majority of the TF-silenced HMDM exhibited significant reductions in *TGFB*, *COL1A1,* and *PDGFRa* (**Figure 4J**). In line with the suppressed pro-fibrotic genes, we observed decreased ⍺SMA by flow cytometry in si*MAZ*, si*EGR1* and a trend in si*KLF3* (**Figure 4K**). In contrast to the other TFs, si*MAFK* conditioned medium increased *TGFB*, *COL1A*2, and *PDGFRa* levels (**Figure 4J**). However, interestingly, the silencing of this transcription factor reduced ⍺SMA levels in fibroblasts.

### Cellular programs and cell-cell communication of macrophages in heart failure

To evaluate the generalizability of the uncovered cellular programs, we first interrogated the relationships of these signatures to a recent study leveraging scRNA-seq data to identify pathogenic macrophages in heart failure [30]. The significant standalone latent factor MD-S-4.1, which our SLIDE model identified to distinguish the transcriptome of macrophages near activated fibroblasts from the ones proximal to resting fibroblasts, is sufficient at providing significant discrimination among cardiac macrophages from the control (Donor), acute myocardial infarction (AMI), and ischemic cardiomyopathy (ICM) groups (**Figure 5A**). The cellular program we identified is higher in ICM and AMI compared to healthy controls. Further, the latent factor is higher in ICM relative to AMI. The ability to extrapolate the identified cellular program to an orthogonal study demonstrates its generalizability.

**Figure 5:**
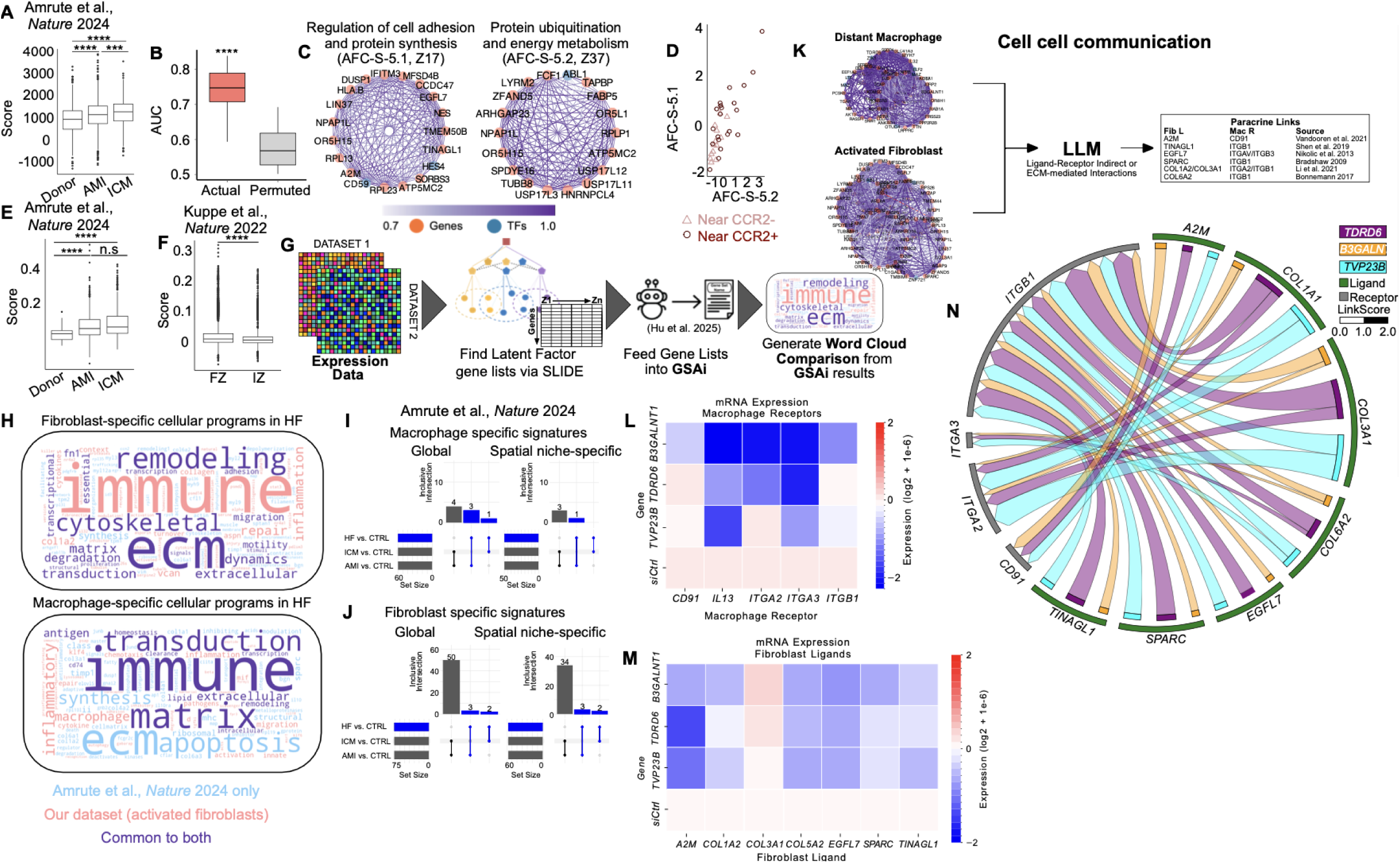
Contextualizing the uncovered cellular programs and unraveling cell-cell communication. A: Overlap of the latent factor M-D-4.1 with the Amrute et al., Nature 2024 data in donor, acute MI, and ischemic cardiomyopathy myeloid cells. B: SLIDE model performance for activated fibroblasts proximal to CCR2 positive myeloid cells in Amrute et al., Nature 2024. C: Significant standalone LFs visualized as correlation networks-regulation of cell adhesion and protein synthesis (AFC-S-5.1) and protein ubiquitination and energy metabolism (AFC-S-5.2). D: Latent factors AFC-S-5.1 and AFC-S-5.2 distinguish activated fibroblasts proximal to CCR2+ macrophages compared to those proximal to CCR2-macrophages, E: Overlap of the latent factor AFC-S-5.1 with the Amrute et al., Nature 2024 data in donor, acute MI, and ischemic cardiomyopathy myeloid cells. F: Overlap of the latent factor AFC-S-5.1 with the Kuppe et al., Nature 2022 data in fibrotic zone (FZ) vs. ischemic zone (IZ). G: Schematic of the re-analysis pipeline (SLIDE followed by re-analysis using GSAI), H: Fibroblast-specific cellular programs in HF discovered in Amrute et al., *Nature* 2024 and the present study, I: Evaluation of the sensitivity and specificity of the inferred macrophage-specific signatures, J: Evaluation of the sensitivity and specificity of the inferred fibroblast-specific signatures, K: Inferred cell-cell communication involving components of the significant latent factors, L: Validation (expression level) of macrophage receptors, M: Validation (expression level) of fibroblast receptors, N: Validated cell-cell communication visualized as a Circos plot.

Next, to more comprehensively elucidate paracrine signaling, we interrogated differences in fibroblast signatures in our dataset based on their proximity to pathogenic CCR2^+^ macrophages. The prior studies identified CCR2^+^ macrophages as the key population to drive pathogenic fibrosis after MI [30, 31]. We identified two significant standalone latent factors and three interacting latent factors (**Figures 5B, 5C, and S7**). Overall, the expression programs captured by just the two significant standalone latent factors provided excellent discrimination between fibroblasts proximal and distal to CCR2^+^ macrophages (**Figure 5D**). Interestingly, these cellular programs were generalizable to the recent study leveraging scRNA-seq data to identify pathogenic macrophages in heart failure (**Figure 5E**). The fibroblast cellular program based on the proximity of CCR2^+^ vs. CCR2^−^ macrophages is higher in ICM and AMI compared to healthy controls (**Figure 5E**). We also evaluated the generalizability of the latent factor to another independent study leveraging spatial transcriptomic data to define differences between different zones of the heart in the context of MI [31]. Here too, the fibroblast latent factor, discriminating the CCR2^+^ vs. CCR2^−^ macrophage niches, was higher in the fibrotic zone relative to the ischemic zone (**Figure 5F**). These findings demonstrate the power of our interpretable machine learning framework in capturing cell type-specific cellular programs that encapsulates spatial microniches reflective of altered paracrine signaling.

Next, we sought to identify cellular programs from these two studies [30, 31] to assess how well they agreed with our findings. Specifically, we re-analyzed data from the two studies using SLIDE to identify differences between AMI and healthy controls (AMI/ICM scRNA-seq study) and the fibrotic and ischemic zones (MI spatial study). We evaluated the concordance of the latent factors identified from our study and those identified from the AMI/ICM scRNA-seq [30] and MI spatial study [31] using a novel pipeline leveraging LLM-based annotation of de-novo gene sets (**Figure 5G**). We found excellent concordance across the 3 studies (**Figure 5H, Figures S8-S9**). However, our analyses have far higher specificity and sensitivity than conventional DEG analyses (**Figures 5I and 5J, Figure S10)**. Specifically, ∼30-50% of latent factor genes are DEGs. However, the other members of the latent factors are significant in a multivariate setting in the context of the overall group, and would be missed by conventional DEG analyses. More importantly, the overall sizes of the latent factors were significantly lower than that of the DEGs, further confirming that these latent factors represent a prioritized necessary and sufficient set of context-specific co-expression modules for the comparisons of interest (**Figures 5I and 5J, Figure S10).**

Finally, to evaluate changes in spatial cell-cell communication, we used an LLM-based approach to identify receptors in the macrophage latent factors and ligands in the fibroblast latent factors that could explain altered paracrine signaling (**Figure 5K**). Changes in the expression for these receptors and ligands were then evaluated experimentally both after control siRNA treatment and upon silencing of the three key genes distinguishing the microniches and prioritized by our analyses (*B3galnt1, Tdrd6,* and *Tvp23b*). There were several significant changes in the expression of both receptors in macrophages (**Figure 5L**) and ligands in fibroblasts (**Figure 5M**). The expression of some of these receptors and ligands were significantly restrained in the si*B3galnt1,* si*Tdrd6,* and si*Tvp23b* groups, suggesting that inhibiting these genes has significant effects on paracrine signaling between macrophages and activated fibroblasts. Overall, these led to a complex network of altered paracrine signaling (**Figure 5N**).

### Dysregulated macrophage spatial microniches contribute to cardiac remodeling after MI

To further mechanistically elucidate the prioritized pathological drivers of MI-induced HF, we evaluated three genes (*B3galnt1, Tdrd6,* and *Tvp23b*) and two transcription factors (*Mafk* and *Maz*) in a murine model of MI (**Figure 6A**). These five hits were chosen based on systematic computational criteria as described in the sections corresponding to Figs 2 and 4 and the *in vitro* data to validate the hits (**Figure 4**). For the initial hits for which the *in vitro* profiling was performed (**Figure 4**), composite scores were calculated based on the PCR and flow cytometry data. Based on the score, the top 5 genes/TFs, for which no prior functional evidence in macrophages in the context of MI/HF were selected for *in vivo* analyses.

**Figure 6:**
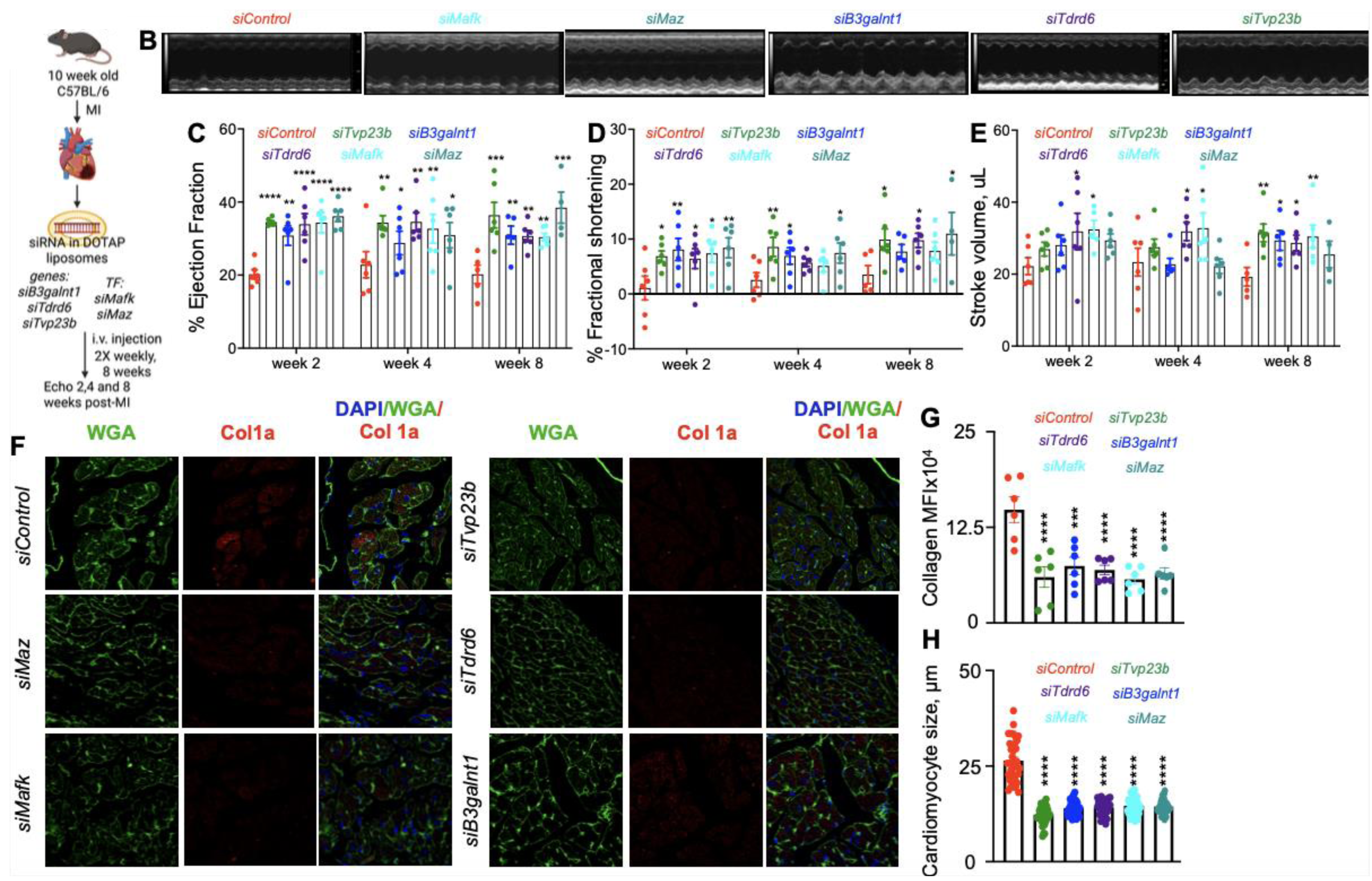
*In vivo* validation of the significance of dysregulated spatial microniches of macrophages in MI-induced HF. A: The schematic shows the experimental design to silence the genes in macrophages after MI. B: Representative images from M mode echocardiography at 28 days after MI are included. C-E: Ejection fraction (C), fractional shortening (D), and stroke volume (E) at weeks 2, 4 and 8 following MI in mice treated with the siRNA were assessed by echocardiography. F-H: Confocal microscopy was performed to quantify Col1a and cardiomyocyte size using WGA staining (F: representative images, G: collagen 1a quantification, and H: cardiomyocyte size quantification). *p<0.05, **p<0.01, ***p<0.001, ****p<0.0001. one-way ANOVA with post-hoc Fisher LSD test. n=5-6/group.

We next validated increased expression of the selected genes and TFs. First, we examined the expression of *B3GALNT1*, *TDRD6*, and *TVP23B* in macrophages proximal to resting and activated fibroblasts. In our spatial data, the expression of these genes was greater in macrophages near activated fibroblasts (**Figure S11**). Further, we observed that cardiac macrophages in MI hearts had higher expression of B3GALNT1, TDRD6, and TVP23B (**Figure S12**). Additionally, we observed increased MAFK but not MAZ in the infarct.

siRNA against the selected genes and transcription factors was formulated in DOTAP, which is specifically engulfed by macrophages [51] and injected in mice after MI. Longitudinal echocardiography was performed at 2, 4, and 8 weeks after MI (**Figure 6A**). Representative images from the echocardiography analyses are shown in **Figure 6B**. Cardiac ejection fraction was significantly higher in mice treated with the latent factor siRNAs *siB3galnt1, siTvp23b,* and *siTdrd6*, and the transcription factor siRNAs *siMafk* and *siMaz* (**Figure 6C**). This beneficial effect on cardiac ejection fraction persisted at weeks 4 and 8 after MI. Fractional shortening, a measure of ventricular shortening during systole, was greater in all experimental groups compared to siControl at 2 weeks following MI (**Figure 6D**). Furthermore, while we observed non-significant increases in stroke volume in the treatment groups compared to the control at weeks 2 and 4, all but the si*Maz* group had significantly improved stroke volume compared to the siControl group at week 8 after MI (**Figure 6E**). Immunostaining analyses of heart sections showed lesser Collagen 1a staining in the experimental groups compared to siControl (**Figure 6F-6G**). Furthermore, we observe reduced cardiomyocyte size in the experimental groups (**Figure 6H**). Interestingly, silencing of the genes and TFs did not significantly alter the abundance of the leukocyte subsets in the blood, bone marrow, and spleen (**Figure S13**).

## Discussion

Inflammation is a pivotal factor in the pathophysiology of heart failure, and elevated levels of inflammatory markers are inversely correlated with clinical outcomes in HF patients [19, 20]. Persistent inflammation exaggerates the recruitment of circulatory monocytes to the heart, wherein they differentiate into proinflammatory macrophages, promoting adverse cardiac remodeling and worsening cardiac function [52, 53]. A complex network of cellular processes and intercellular communications underlie cardiac homeostasis. While single-cell transcriptomics and multi-omics approaches have shed light on disruptions in the transcriptional and regulatory networks of cardiac cell types in HF, they fail to capture complex paracrine signaling and intercellular interactions. Here, we demonstrate comprehensive spatial analyses of cardiac macrophages, resting fibroblasts, and activated fibroblasts in ischemic cardiomyopathy. Using a novel framework coupling interpretable machine learning, regulatory network analyses and in-silico perturbations, we demonstrate differences in the transcriptomic and gene regulatory state of cardiac macrophages proximal and distal to activated fibroblasts. Thus far, spatial transcriptomics studies of human hearts and ischemic heart failure have described the transcriptomic differences in different cell types in the heart, and intercellular communication is deciphered via computational approaches [31, 32, 54–56]. However, these studies do not provide the transcriptomic profiles of macrophages based on their location in respect to fibroblast subsets. Here, we provide a comprehensive map of cardiac fibroblasts, macrophage dynamics, and identify microniche-specific cellular programs in macrophages that likely drive pathogenic processes in ischemic cardiomyopathy. For example, epigenetic reader methyl-CpG-binding domain protein 2 (MBD2) is known to regulate macrophage polarization and play a pro-fibrotic role in bleomycin-induced pulmonary fibrosis and renal fibrosis [37, 38]. Adipocyte Enhancer Binding Protein 1 (AEBP1) is a known regulator of renal, liver, and cardiac fibrosis. Studies have explored the therapeutic potential of AEBP1 inhibition in heart failure [39, 40].

A key conceptual innovation of the study involves moving beyond conventional DE analyses as these are underpowered to pick up nuanced differences or hits beyond what has already been discovered. For example, DE analyses comparing HF and control samples only uncovered known fibrotic markers while our machine learning analyses identified several novel hits, which were functionally validated. More importantly, complex spatial microniche-specific discriminators between macrophages proximal to activated or resting fibroblasts were entirely missed by DE analyses. Our systems framework identified these discriminators, and the functional importance of the identified hits was validated using *in vitro* and *in vivo* analyses. Critically, for the in-vivo analyses, we evaluated three genes (*B3galnt1, Tdrd6,* and *Tvp23b*) and two transcription factors (*Mafk* and *Maz*) in a murine model of MI. All 5 had significant phenotypes in-vivo in terms of multiple readouts of cardiac function, demonstrating the ability of our approach to identify functionally meaningful hits. Our signatures were also replicated across orthogonal cohorts [30, 31]. Another critical aspect that enabled meaningful functional validation was the interpretability of the machine learning approaches. While deep learning approaches are extremely powerful at prediction, they are often uninterpretable black boxes and cannot be used effectively for meaningful inference and downstream functional validation. Together, these findings suggest that the multivariate interpretable machine learning framework we developed here can be broadly extrapolated across complex disease contexts and is well suited for functional discovery of mechanistic drivers.

We prioritized *TDRD6*, *B3GALNT1*, *TVP23B*, *IQGAP1*, *AASDHPPT*, *GPR107*, and *SERBP1* in human monocyte-derived macrophages and cardiac fibroblasts based on the same systematic criteria described above (contributions to these significant latent factors and individual effect sizes). The functions of most of these genes in the context of cardiovascular disease are not well known. TDRD6 is a tudor domain containing protein and is important for chromatid body formation during spermatogenesis [57]. B3GALNT1 is a member of the beta-1,3-galactosyltransferase (beta3GalT) gene family, and it is a possible therapeutic target for non-small cell lung cancer [58]. TVP23B and GPR107 localize to the Golgi apparatus and have potential roles in vesicular trafficking, intestinal homeostasis, receptor-mediated endocytosis, and Ldlr uptake [59–61]. IQGAP1 is a scaffolding protein that has cellular functions including cytoskeletal dynamics, vesicular trafficking, MAPK signaling, cell migration, and cancer metastasis [62–64]. AASDHPPT localizes to mitochondria and catalyzes the post translational modification of target proteins by transferring phosphopantetheine from CoA. This gene is required for mitochondrial respiration, and loss of AASDHPPT diminishes mitochondrial fatty acid synthesis [65]. SERBP1 is an RNA-binding protein and regulates glioblastoma multiforme development [66]. Based on the effects of the silencing of these genes in vitro, we chose *TDRD6*, *B3GALNT1,* and *TVP23B* for further *in vivo* validation. Additionally, we discerned macrophage-specific functions of *MAFK* and *MAZ*, which are TFs, in mice with MI. Macrophage-specific downregulation of these genes and TFs using a lipidoid nanoparticle technique significantly improved cardiac function and attenuated cardiac fibrosis. Future clinical trials will be required to evaluate the significance of these genes and TFs in patients with STEMI.

Our study has several limitations. We utilized myocardial specimens from ischemic heart failure patients who underwent heart transplant i.e the samples were collected at the advanced stage of the disease. Therefore, although we uncovered novel insights with our analyses, we do not have a temporal insight into myeloid – fibroblast signaling in heart failure. Additionally, silencing *TDRD6*, *B3GALNT1,* and *TVP23B* increased fibrotic gene expression in macrophages *in vitro*; however, we observed an improvement in cardiac function and remodeling in mice with MI after silencing these genes specifically in macrophages *in vivo*. This observation is in line with the finding by Amrute et al. that cultured fibroblasts sub-optimally capture cardiac injury [30]. Furthermore, while we focused on macrophages, other immune cell types like T cells shape the inflammatory niche of the failing heart. Future studies are required to obtain further insights at the temporal signaling events and transcriptomic signature of other immune cell types, endothelial cells, and cardiomyocytes in heart failure. Finally, an LLM-based approach uncovered several receptors in the macrophage latent factors and ligands in the fibroblast latent factors that could explain altered paracrine signaling between these cell types. Elucidating the significance of these ligand-receptor pairs in MI is beyond the scope of this manuscript. Future mechanistic studies using mice with macrophage and fibroblast-specific deficiency of these receptor-ligand pairs will be required to understand their functions in post-MI cardiac remodeling.

## Acknowledgements

This work benefitted from Cytek Aurora CS Spectral Sorter funded by NIH S10OD032265 and Olympus FV3000 confocal microscope funded by NIH S10OD030254. This research was funded by the National Institutes of Health (NIH) grants R00HL121076, R01HL142629, R01HL142629-04W1, R01HL143967, R01AG069399 and R01DK129339, the VA Merit Award I01BX006392-01, the American Heart Association (AHA) Transformational Project Award (19TPA34910142), the AHA Innovative Project Award (19IPLOI34760566), the AHA Established Project Award (25EIA14115660), the AHA Strategically Focused Research Network Award (24SFRNPCN1280229), the ALA Innovation Project Award (IA-629694), and the AHA Innovative Project Award (23IPA1053549) to P.D; NHLBI K99/R00HL157689 to NN; DP2AI164325, U01HG012041, R01AI170108, U01AI17951 to JD.

## Methods

Our research complies with all relevant ethical regulations and were approved by the University of Pittsburgh Institutional Review Board (IRB approvals: STUDY19090164), Institutional Animal Care and Use Committee.

### Human cardiac tissue

Human left ventricle specimens from transplant recipients with ischemic cardiomyopathy were obtained through a collaboration with Dr. Charles McTiernan (IRB approvals: STUDY19090164). These patients had history of MI. Heart tissues were obtained from patients after informed consent at University of Pittsburgh Medical Center. No compensation was provided for patient participation in this study. Demographic information of heart failure patient and controls is de-identified and summarized in **Figure S1A**. Immune status, medical history and medications / drug usage data are unknown. Control human heart tissues were obtained through a collaboration with Dr. John Sembrat. The control patients had no known history of cardiac disease and died due to non-cardiac causes. All samples were preserved in cold cardioplegic solution for transport to the laboratory. The specimens were then fixed in 4% formaldehyde for 24 hours and embedded in OCT compound. OCT blocks were sectioned (10 μm) for GeoMx analyses. After sectioning, the sections were stained and processed according to manufacturer’s protocol. Briefly, 10 μm sections were obtained and stained with CD68 (macrophage), vimentin (resting fibroblasts) and alpha smooth muscle actin (activated fibroblasts). Roughly 10 areas were captured per specimen for analysis.

### GeoMX Pre-Processing

Raw Digital Count Conversion (DCC) files containing expression count data and sequencing quality metadata were processed using the GeomxTools v.3.8.0, and NanoStringNCTools v.1.12.0 packages. Probe assay metadata was imported from probe kit configuration (PKC) files, and sample annotations were incorporated from external spreadsheets.

Sequencing quality control (QC) was applied per region of interest (ROI) using shiftCountsOne and setSegmentQCFlags. ROIs were filtered based on minimum thresholds for sequencing reads (1,000), trimming (≥80%), stitching (≥80%), alignment (≥75%), and saturation (≥50%), along with sample-specific metrics such as minimum nuclei (≥20), area (≥1,000 µm²), and background controls (minimum negative counts ≥5; NTC maximum ≤9,000). Segments were flagged for quality and retained if they passed all QC criteria. Probe-level QC was conducted using setBioProbeQCFlags, removing local and global outliers based on Grubbs’ test and requiring a minimum probe ratio of 0.1 and failure rates <20%. Region-level expression counts were aggregated to gene-level using aggregateCounts. The limit of quantification (LOQ) was calculated per segment and per module using the geometric mean and standard deviation of negative probes. Genes were considered detected if expression exceeded the module-specific LOQ. Gene filtering was performed by computing per-gene detection rates across ROIs. Genes were retained if detected in ≥3% of segments or classified as negative controls. Finally, quantile normalization (75th percentile) was applied using normalize with the “quant” method, storing normalized counts in the q_norm expression slot. The preprocessed and normalized data were used for subsequent downstream analyses of spatial gene expression patterns.

### GeoMX UMAP

Dimensionality reduction and visualization of the GeoMX data were performed using Scanpy package. Following normalization and quality control, we first created an AnnData object from the normalized expression data and raw count data. Sample metadata, including segment labels and disease status (healthy vs. heart failure), were integrated from an external annotation file. Prior to dimensionality reduction, the expression data underwent mean centering to standardize gene expression values across samples. Principal component analysis (PCA) was then applied for initial dimensionality reduction, followed by Louvain community detection for unsupervised clustering. Uniform Manifold Approximation and Projection (UMAP) was subsequently performed to visualize the high-dimensional data in a two-dimensional space, enabling identification of cell population structure and relationships. To investigate cell type-specific differences between disease states, we created a multi-panel UMAP visualization where each spatial segment type was plotted separately. For each cell type, a separate UMAP plot was generated, with points colored according to disease status. All visualizations were created using Scanpy’s plotting functions with standardized point sizes to ensure visibility.

### DESeq2 Analysis

Differential expression analysis was performed using DESeq2 v.1.44.0 in R to identify genes with significant expression changes between conditions. A DESeqDataSet object was constructed using the count matrix and condition information, with the design formula specifying condition as the variable of interest. Differential expression analysis was conducted using the DESeq function with default parameters, except for disabling the beta prior (betaPrior = FALSE) for log fold change estimation. To correct for the high variance of log fold change estimates for low-count genes, we applied the lfcShrink function using the coefficient for the second level of the condition factor. Differentially expressed genes were visualized using a custom implementation of the EnhancedVolcano v.1.22.0 package. The volcano plot displayed the relationship between statistical significance (-log10 of adjusted p-values) and the magnitude of change (log2 fold change). Genes were categorized into four groups: (1) not significant, (2) significant by log2 fold change only, (3) significant by adjusted p-value only, and (4) significant by both criteria. Significance thresholds were set at an adjusted p-value of 10e-6 and a log2 fold change of ±1.

### SLIDE Models

To identify key latent factors and their interactions underlying conditions of interest, we employed SLIDE, a supervised, identifiable, data-distribution-free approach for analyzing high-dimensional datasets. Unlike traditional methods that often require assumptions about data distribution or rely on prior biological knowledge, SLIDE implements a latent-factor regression framework that can detect both linear and nonlinear relationships, including complex hierarchical structures, within the data. SLIDE operates through a two-step process. First, the latent factor discovery framework which performs dimensionality reduction to transform observable features (Xs) into latent factors (Zs) with stringent identifiability guarantees. These latent factors were then regressed against outcomes of interest while maintaining stringent false discovery rate (FDR) control. For all analyses, we maintained a type I error threshold (alpha) of 0.05. SLIDE integrates a modified knockoff technique for this FDR control, providing formal statistical guarantees that the discovered latent factors represent genuine biological signals rather than statistical artifacts.

SLIDE explores an extensive search space of potential relationships to identify a small subset of biologically relevant latent factors (gene modules) that significantly influence the phenotype of interest. Critically, it incorporates interaction terms between latent factors to capture complex regulatory relationships that may not be evident when examining individual factors in isolation. To ensure that final models were not overfit, we implemented nested k-fold cross-validation with permutation testing, further strengthening the reliability of our findings.

### SLIDE models to discriminate cell-type specific HF and Control regions

To uncover cell-type latent factors that could stratify MI and Control regions, we utilized the normalized counts of active fibroblasts, resting fibroblasts, and macrophages. The SLIDE models involve tuning two key hyperparameters: delta and lambda. Delta determines the number of latent factors extracted from the data, while lambda controls the sparsity of each latent factor. Hyperparameter optimization was performed using a grid search method embedded in the SLIDE framework. To ensure robust model performance, we implemented a rigorous multi-replicate 10-fold cross-validation approach. The regions were randomly divided into 10 equal subsets, with 9 subsets used as training data and 1 subset used as testing data in each iteration. This process was repeated until each subset had served as the test set once for multiple replicates, providing comprehensive validation of the model’s predictive capability across the entire dataset.

### SLIDE Models to discriminate regions with distinct local environment and localization

To discover distinct cellular programs in macrophages with different proximity to activated fibroblast, we used the nuclei counts of active fibroblasts, resting fibroblasts, and macrophages from each region across all tissues in HF. To classify regions based on fibroblast activation status, we calculated the relative abundance of resting and active fibroblasts within each ROI. We computed normalization ratios by dividing the nuclei count of resting and active fibroblasts in each ROI by their respective total counts across all samples. This normalization accounted for potential differences in overall cell abundance between phenotypes. ROIs were classified into 2 classes based on the predominance of either resting or active fibroblasts: regions where the normalized resting fibroblast ratio exceeded the active fibroblast ratio, and regions with higher active fibroblast ratios. To discover latent factors stratifying activated fibroblasts associated with either CCR2-high or CCR2-low expressing macrophages from HF samples, we divided the regions into 2 classes based on their CCR2 expression profiles. CCR2 expression was quantified in these regions, and a threshold of 10 (raw count value) was used to classify regions as CCR2-low or CCR2-high. Active fibroblast segments were then classified based on their spatial location relative to CCR2-low or CCR2-high macrophage regions.

### SLIDE models for public datasets

For comparative analysis to our data, we utilized publicly available datasets, Kuppe et al. 2023 and Amrute et al. 2024, available at cellxgene https://cellxgene.cziscience.com/ collections/8191c283-0816-424b-9b61-c3e1d6258a77 and the Gene Expres-sion Omnibus super series (GSE218392), respectively. snRNA-seq data were analyzed using the Seurat package in R. Cells were filtered to exclude low quality and doublet values. Quality control was performed to keep nuclei within the following: RNA counts >500 and <25,000, feature counts >500 and <6,000, and percent mito <15%. Filtered raw RNA counts were normalized using NormalizeData default settings. Highly variable features were identified using FindVariableFeatures, selecting the top 2,000 variable genes and the 10 most highly variable genes were extracted using VariableFeatures. Gene expression values were then scaled before performing PCA, nearest neighbor clustering, and UMAP embedding construction.

Latent factor gene sets were produced from SLIDE analysis of publicly available datasets, Kuppe et al. 2023 and Amrute et al. 2024. Raw and processed snRNA-seq and spatial transcriptomics data are available at cellxgene https://cellxgene.cziscience.com/collections/8191c283-0816-424b-9b61-c3e1d6258a77 and the Gene Expression Omnibus super series (GSE218392), respectively. To interpret our latent factors, the gene sets were run through Gene Set AI (GSAi), an LLM for gene set analysis. WordClouds were then generated in Python by comparing GSAi results between groups of interest, to capture shared patterns in output.

To visualize comparative analysis of latent factors, we designed upset plots in R for inclusive DEGs across various groups. Plots were produced following the ComplexUpset protocol described by Michał Krassowski. (2020). krassowski/complex-upset.

### Correlation Network Analysis of Latent Factors

To visualize the relationships between genes within latent factors identified by SLIDE, we constructed correlation networks based on gene expression data. Expression data were imported from a CSV file containing the normalized gene expression matrix. Significant genes associated with each latent factor were loaded from the SLIDE analysis results.

For each identified latent factor, a correlation network was constructed using the following procedure. First, the subset of genes belonging to the specific latent factor was extracted from the expression matrix, and a pairwise Pearson correlation matrix was calculated using the cor function in R. Network visualization was performed using the Cytoscape, which enables the representation of correlation matrices as networks where genes are depicted as nodes and correlations as edges. To emphasize meaningful relationships, a correlation threshold of 0.7 was applied, whereby only correlations exceeding this absolute value were visualized as edges.

### Cell Type-Specific Gene Regulatory Network Reconstruction

We adapted CellOracle, a computational framework designed for scRNA-seq data for context-specific gene regulatory network (GRN) inference and in silico perturbation, to investigate transcriptional regulation in out GeoMX dataset. First, normalized expression data from each cell type were imported and quality-filtered, with sample metadata integrated from clinical annotations to distinguish between HF and control conditions. Since CellOracle is designed for scRNA-seq datasets with higher feature dimensions, we adjusted the quality control code embedded in CellOracle to allow a smaller number of genes than it’s default at 999 genes.

Dimensionality reduction was performed through sequential mean centering, principal component analysis (PCA), and uniform manifold approximation and projection (UMAP) to visualize the transcriptional landscape. Cell clustering was implemented using both Leiden and Louvain community detection algorithms with a neighborhood graph constructed from 50 principal components and 5 nearest neighbors. The resulting two-dimensional embeddings revealed distinct transcriptional states within the cell population that were visualized with respect to disease status (HF vs. control) and cluster assignment.

### Context-Specific Regulatory Network Inference

A base regulatory network was established using the human promoter-based GRN available in CellOracle, which contains documented transcription factor (TF) binding site information. Context-specific, cluster-specific gene regulatory relationships were inferred using the aggregated data from cell clusters identified by the Louvain algorithm. The GRN inference was performed with a regularization parameter (alpha) of 10 to control for model complexity. The resulting links between TFs and target genes were filtered based on statistical significance (p-value) and further prioritized using a weighted log-p metric that combines the coefficient magnitude and statistical significance.

### Latent Factors-GRN integration for Context-Specific Perturbation Target Prioritization

The top 10% of regulatory links were retained based on their weighted significance scores, and the regulatory influence of each TF was quantified through its node degree within the network. To identify biologically meaningful regulatory relationships, we integrated our GRN analysis with results from SLIDE analysis, which had identified latent factors associated with MI or micro-environment in our dataset. We identified TFs represented within these latent factors by evaluating their overlap with known regulatory genes curated from a context-specific GRN. For each regulatory cluster in the GRN, we computed the intersection between the set of genes in the latent factors and the set of TFs active within that cluster. This allowed us to determine which latent factors potentially reflect underlying transcriptional regulation.

In parallel, we investigated the TFs that may regulate the genes within each latent factor. For each cluster-specific GRN, we selected edges in which the target genes were present in a latent factor and the source nodes were TFs. This enabled us to link TFs to latent factor genes based on prior knowledge of regulatory interactions. The resulting associations between TFs and latent factor genes were used to identify key regulators and assess their connectivity within each cluster. To estimate the regulatory influence of these TFs quantitatively, we applied regularized linear regression using TF activity profiles as predictors and gene expression as response variables. This yielded a weighted, context-specific GRN reflecting the influence of TFs on latent gene modules across spatial contexts.

### In Silico Transcription Factor Perturbation

To predict the functional impact of key transcription factors on macrophage state transitions, we performed in silico perturbation experiments using CellOracle’s vector field-based approach. TFs were selected for perturbation based on their presence in SLIDE-derived latent factors or their high regulatory connectivity to latent factor genes (threshold connectivity ≥ 8).

For each selected TF, we first examined its expression distribution across the macrophage population and then simulated complete loss-of-function (setting expression to 0.0) to predict cellular identity shifts. The resultant transcriptional trajectories were visualized as vector fields overlaid on the UMAP embedding, with a scale factor of 15 applied to enhance visualization of predicted cell state transitions.

The optimal minimum mass parameter for vector field calculation was determined through systematic grid search across 40 parameter values. Cluster-specific effects of TF perturbations were quantified by calculating vector field differences between baseline and perturbed conditions for each Louvain cluster. This analysis enabled identification of macrophage subpopulations most susceptible to transcriptional reprogramming upon perturbation of heart failure-associated regulatory factors.

### siRNA Pairwise SVM Classification Analysis

To evaluate the discriminative power of our siRNA targets compared to non-targeting (NT) controls, we implemented a binary classification approach using Support Vector Machines (SVM). This analysis was performed separately on three different data types: qPCR results, flow cytometry frequency measurements, and flow cytometry Mean Fluorescence Intensity (MFI) values. We developed a cross-validation framework to assess the classification performance between each siRNA target and the NT control. For each target pair (a specific siRNA target vs. NT control), we separated the relevant samples from the complete dataset and encoded the target labels into binary numerical values. We implemented a stratified k-fold cross-validation (k=4) with random shuffling to ensure consistent fold distributions across experiments. Within each fold, an SVM classifier with probability estimation (SVC with probability=True) was trained on the training subset, after which the model predicted class probabilities for the test subset. The area under the receiver operating characteristic curve (AUC) was calculated using the predicted probabilities and true labels. To ensure robust performance estimates, this entire cross-validation procedure was repeated 100 times for each target pair. The mean AUC across all folds and iterations was calculated for each target pair, serving as our primary metric for classification performance.

### Transcription Factor Similarity Analysis

We first categorized targets as either TFs or non-TFs by cross-referencing our target list with a comprehensive database of human TFs. We quantified the similarity between expression profiles using cosine similarity, which measures the cosine of the angle between two non-zero vectors in a multi-dimensional space. For each TF and non-TF pair, we calculated the cosine similarity between their respective feature vectors using the formula:

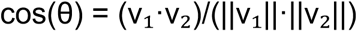

where v₁ and v₂ represent the feature vectors of two samples, · denotes the dot product, and ||v|| represents the Euclidean norm of vector v.

We performed this calculation between each sample of a given TF and each sample of a given non-TF, resulting in a comprehensive set of pairwise similarity scores. We then computed the mean similarity score for each TF-gene pair to obtain a representative measure of their overall similarity.

The resulting similarity matrices were visualized as heatmaps using Plotly, with color intensity representing the degree of similarity (scaled from 0.7 to 1.0 using a blue color gradient). This visualization approach allowed for intuitive identification of highly similar TF and non-TF pairs. We generated separate heatmaps for: TF vs. non-TF similarities, and TF vs. control (NT and TGFβ) similarities.

### Monocyte-derived macrophage culture and gene silencing

Leukopaks were obtained from StemCell technologies. Following RBC lysis and platelet removal, PBMCs were isolated by centrifugation in LymphoprepTM density gradient medium (Stemcell technologies). Monocytes were isolated from PBMC using CD14 enrichment kit without CD16 depletion (Stemcell technologies) and cultured in the presence of 50 ng/ml M-CSF for 7 days to differentiate them into macrophages. Gene silencing was performed with Lipofectamine RNAiMax (Life Technologies). MDMs were treated with 100ug/ml oxLDL for 48 hours starting at 24 hours after gene silencing. Conditioned medium was harvested for cardiac fibroblast culture. RNeasy RNA isolation kit (Qiagen) was used to extract RNA and cDNA was synthesized using Applied Biosystems High Capacity RNA to cDNA kit. Gene expression was quantified by quantitative RT-PCR using SYBR green mastermix, and primers (IDT). Gene expression Ct values were normalized to those of β-actin.

### Human cardiac fibroblasts

Human cardiac left ventricular fibroblasts were purchased from Lonza (CC-2904) and maintained in complete fibroblast medium (Lonza CC-4526). Cardiac fibroblasts were treated with 20% by volume conditioned medium from MDMs for 48 hours. Flow cytometry was performed to quantify fibroblast differentiation by aSMA staining.

### Animals

All animal work was approved by the University of Pittsburgh IACUC committee. The experiments were carried out in 10-week-old female C57BL/6 mice were randomized into 6 groups – *siControl, siB3galnt1, siTvp23b, siTdrd6, siMafk,* and *siMaz*.

#### Myocardial infarction

MI was induced in 10-week-old female C57BL/6 mice by LAD ligation as previously described [67]. Briefly, animals were anesthetized, intubated and maintained at a heart rate of 137 beats per minute and a tidal volume of 0.18 cc. An incision was made in the third intercostal space, the ribs were spread using retractors to expose the left ventricle. The left coronary artery was ligated using a 8-0 nylon suture with the help of a needle holder. Successful ligation was confirmed by visualizing the blanching of the lower left ventricle. The retractors were removed, and the skin was sutured with 4-0 nylon suture. Animals were maintained under a heat lamp for recovery.

### In vivo gene silencing with DOTAP

siRNAs were administered intravenously in DOTAP liposomes twice a week for 8 weeks of the experiment. DOTAP:cholesterol liposomes (Encapsula Nanosciences) were administered at 1:7.5 siRNA:total DOTAP by weight and a dosage of 1.5mg/ kg DOTAP. Echocardiography was performed at 14 days, 28 days, and 8 weeks following MI.

### Organ harvesting and flow cytometry

Mice were euthanized 8 weeks after MI, according to the University of Pittsburgh IACUC guidelines followed by perfusion of PBS through the left ventricle. Blood was collected by terminal cardiac puncture and incubated with RBC lysis buffer for 3 minutes at room temperature, followed by addition of FACS buffer and centrifugation to pellet leukocytes. A hemocytometer was used to count the number of viable cells in the organs. The following panel of antibodies were used to analyze the myeloid cell populations in mice: anti-CD45.2 (Biolegend 109820, clone 104), CD11b (BD Biosciences 557657, clone M1/70), CD115 (Biolegend 565249, clone T38-320), Ly6G (Biolegend 563979, clone 1A8), Ly-6C (eBioscience 45-5932-82, clone HK1.4), F4/80 (Biolegend 123114, clone BM8) CD3 (BD Biosciences 367-0032-82, clone 17A2), and CD19 (BD Biosciences 563148, clone 1D3). Neutrophils were identified as CD45^+^, CD11b^+^, Ly6G^+^, and CD115^−^. Monocytes were considered as CD45^+^, CD11b^+^, Ly-6G^−^, and CD115^+^. B and T lymphocytes were identified as CD45^+^, CD11b^−^, CD19^+^, CD3^−^ and CD45^+^, CD11b^−^, CD19^−^, CD3^+^, respectively. A Cytek Aurora flow cytometer was used to acquire the data, which were analyzed with the FlowJo software (Tree Star).

### Immunofluorescence

Sham and MI hearts were harvested 4 weeks post-MI and embedded in OCT blocks. 10 μm sections were obtained for immunostaining (B3galnt1, Tvp23b, Tdrd6, Mafk, and Maz). Hearts from MI animals treated with siRNA-DOTAP were harvested, fixed overnight in 4% formaldehyde, and embedded in paraffin blocks. 5 μm sections were obtained from these blocks for immunostaining (Collagen and wheat germ agglutinin). Briefly, deparaffinization and rehydration of FFPE sections was carried out with xylene and ethanol. After blocking with 5% BSA, sections were incubated with primary antibodies overnight, washed, and incubated with secondary fluorescent antibodies for an hour. Sections were imaged with an Olympus confocal microscope.

### Statistical analysis

Data are displayed as mean ± SEM. Statistical significance between groups was performed using Graphpad PRISM. A two-tailed Student’s t-test was used to compare the means of two groups. One-way ANOVA with post-hoc Fisher LSD test was used to compare >2 groups. Results were considered as statistically significant when p < 0.05.

## Data Availability

The spatial transcriptomic data generated in this study have been deposited to GSE297540. They can be accessed using the reviewer token mxqpuuokvvwdvkl.

## Code Availability

Code and documentation for all the analyses in the manuscript is available at https://github.com/Hanxi-002/MI_Spatial.

## Supplementary figure legends

**Figure S1 – Study cohort details (Accompanies Figure 1)**

A: Summary of ischemic cardiomyopathy and control patient demographics. B: Bar plot showing the gene detection rate across samples in the GeoMX dataset. Samples were categorized by detection rate thresholds: <1%, 1–5%, 5–10%, 10–15%, and >15%, with height corresponding to the proportion of genes detected above the calculated limit of quantitation (LOQ). Detection rate is defined as the fraction of targets with expression above LOQ.

**Figure S2 – SLIDE analyses of HF and control macrophages (Accompanies Figure 2).**

A: Significant latent factors identified by SLIDE stratifying HF and control macrophages. Blue circles represent standalone latent factors, and coral circles represent interacting latent factors. B-D: Correlation networks of gene co-expression for the three interacting latent factors. The latent factors are shown as M-I-E-2.1 (Z4) (B), M-I-E-2.2 (Z6) (C), and M-I-E-2.3 (Z22) (D). The green nodes represent genes, and the yellow nodes represent transcription factors. The color gradient of the connecting edges indicates the strength of correlation among the connected genes. E: Validation of target gene silencing with siRNA. N=4/group, * *p*<0.05, ** *p*<0.01, 2-tailed Student’s t-test F: Pro-fibrotic gene expression in bone marrow-derived macrophages after silencing of *Aebp1*, *Ciita* and *Mbd2*. N=4/group, * *p*<0.05, ** *p*<0.01, *** p<0.001, **** p<0.0001. one-way ANOVA with post-hoc Fisher LSD test. Expression normalized to siControl (dotted red line). G: Fibrotic gene expression in mouse cardiac fibroblasts treated with conditioned medium from *siAebp1*, *siCiita* and *siMbd2*-treated macrophages. N=4/group, * *p*<0.05, ** *p*<0.01, *** p<0.001, **** p<0.0001. one-way ANOVA with post-hoc Fisher LSD test. Expression normalized to siControl (dotted red line). H: Key transcription factors controlling macrophage cellular programs in Control.

**Figure S3 – SLIDE analyses of cardiac fibroblasts in HF and control specimens (Accompanies Figure 3).**

A: Significant latent factors identified by SLIDE stratifying HF and control fibroblasts. The blue circle represents standalone latent factors, and coral circles represent interacting latent factors. B-D: Correlation networks of gene co-expression for the three interacting latent factors. The latent factors are shown as F-I-E-3.1 (Z1) (B), F-I-E-3.2 (Z3) (C), and F-I-E-3.3 (Z4) (D). Green nodes represent genes, and yellow nodes represent transcription factors. The color gradient of the connecting edges indicates the strength of correlation among the connected genes.

**Figure S4 – SLIDE analyses of resting fibroblasts (Accompanies Figure 3).**

A: Volcano plot of differentially expressed genes in HF resting fibroblasts compared to control resting fibroblasts. B: SLIDE model discriminates HF and control resting fibroblasts. C: Significant latent factors identified by SLIDE stratifying HFI and control resting fibroblasts. Blue circles represent standalone latent factors, and coral circles represent interacting latent factors. D-K: Correlation networks of gene co-expression for the thre standalone and four interacting latent factors. The latent factors are shown as RF-S-E-4.1 (Z2) (D), RF-S-E-4.2 (Z18) (E), RF-S-E-4.3 (Z22) (F), RF-I-E-4.1 (Z4) (G), RF-I-E-4.2 (Z5) (H), RF-I-E-4.3 (Z16) (I), RF-I-E-4.4 (Z19) (J), and RF-I-E-4.5 (Z20) (K).The green nodes represent genes, and the yellow nodes represent transcription factors. The color gradient of the connecting edges indicates the strength of correlation among the connected genes.

**Figure S5 – SLIDE analyses of activated fibroblasts in heart failure (Accompanies Figure 3).**

A: Significant latent factors identified by SLIDE stratifying HF and control activated fibroblasts. Blue circles represent standalone latent factors, and coral circles represent interacting latent factors. B-F: Correlation networks of gene co-expression for the five interacting latent factors. The latent factors are shown as AF-I-1 (Z10) (B), AF-I-2 (Z16) (C), AF-I-3 (Z30) (D), AF-I-4 (Z37) (E), and AF-I-5 (Z43) (F). The green nodes represent genes, and the yellow nodes represent transcription factors. The color gradient of the connecting edges indicates the strength of correlation among the connected genes. G: Key TFs controlling cellular programs in control activated fibroblasts filtered based on a high-quality GRN, enrichment in LFs, and in-silico perturbations.

**Figure S6 – SLIDE analyses of macrophages proximal to activated fibroblasts (Accompanies Figure 4).**

A: Significant latent factors identified by SLIDE stratifying HF macrophages in spatial microniches defined by proximity to activated or resting fibroblasts. The green nodes represent genes, and the yellow nodes represent transcription factors. The color gradient of the connecting edges indicates the strength of correlation among the connected genes. The latent factors are shown as MD-I-E-6.1 (Z45) and MD-I-E-6.2 (Z131) (B). The green nodes represent genes, and the yellow nodes represent transcription factors. The color gradient of the connecting edges indicates the strength of correlation among the connected genes. C: Validation of target gene silencing with siRNA in human monocyte-derived macrophages. N=4/group, * *p*<0.05, ** *p*<0.01, *** p<0.001, 2-tailed Student’s t-test D: Key TFs controlling cellular programs in control macrophages filtered based on a high-quality GRN enrichment in LFs and *in silico* perturbations. E: Validation of target transcription factor silencing with siRNA in human monocyte-derived macrophages. N=4/group, * *p*<0.05, ** *p*<0.01, *** p<0.001, 2-tailed Student’s t-test

**Figure S7 – Spatial microniches of fibroblasts proximal to CCR2^+^ or CCR2^−^ macrophages (Accompanies Figure 5).**

A: Significant latent factors identified by SLIDE stratifying HF activated fibroblasts by spatial microniches defined by proximity to macrophages expressing not expressing CCR2.The blue circles represent standalone latent factors, and the coral circles represent interacting latent factors. B: Correlation networks of gene co-expression for the three interacting latent factors. The latent factors are shown as AFC-I-E-7.1 (Z3), AFC-I-E-7.2 (Z21), and AFC-I-E-7.3 (Z27). The orange nodes represent genes, and the blue nodes represent transcription factors. The color gradient of the connecting edges indicates the strength of correlation among the connected genes.

**Figure S8 – Unique and shared biological characteristics of latent factors across our dataset and the open-source dataset (Kuppe et al., *Nature* 2022) (Accompanies Figure 5).**

Word Cloud representations of unique (red or blue) and common (purple) biological characteristics for latent factors from a publicly available dataset (Kuppe et al. 2022) vs. our latent factors run through GSAi. (A) BZ-CTRL (border zone) fibroblasts versus our activated fibroblast latent factors (top) or our CCR2 fibroblast latent factors (bottom). (B) IZ-CTRL (ischemic zone) fibroblasts versus our activated fibroblast latent factors (top) or our CCR2 fibroblast latent factors (bottom).

**Figure S9 – Unique and shared biological characteristics of latent factors across our dataset and the open-source dataset (Amrute et al., *Nature* 2024) (Accompanies Figure 5).**

Word Cloud representations of unique (red or blue) and common (purple) biological characteristics for latent factors from a publicly available dataset (Amrute et al. 2024) vs. our latent factors run through GSAi. (A) top: Our activated fibroblast latent factors and AMI-CTRL (acute myocardial infarction) fibroblasts, middle: Our CCR2+ macrophage-fibroblast latent factors and ICM-CTRL (ischemic cardiomyopathies) fibroblasts, bottom: Our CCR2+ macrophage-fibroblast latent factors and AMI-CTRL (ischemic cardiomyopathies) fibroblasts. (B) top: Our macrophage latent factors and AMI-CTRL (acute myocardial infarction) myeloid cells, middle: Our macrophage spatial niche-specific latent factors and ICM-CTRL (ischemic cardiomyopathies) myeloid cells, bottom: Our macrophage spatial niche-specific latent factors and AMI-CTRL (ischemic cardiomyopathies) myeloid cells.

**Figure S10 – Comparison of latent factor gene set to the opensource datasets (Amrute et al. *Nature* 2024 and Kuppe et al., *Nature* 2022) (Accompanies Figure 5)**

Latent factor gene set inclusive intersections, comparing our gene sets (blue) to (A) Amrute et al. Nature 2024 and (B) Kuppe et al., Nature 2022 gene sets; top: size-matched top DEGs (ranked by log2FC) to the number of unique genes in all latent factors; bottom: full gene sets. (A) Comparing ICM-CTRL and AMI-CTRL CCR2+ macrophage latent factors to (left) our macrophage latent factors and (right) our distant macrophage latent factors. (B) Comparing IZ-CTRL and BZ-CTRL fibroblast latent factors to (left) our activated fibroblast latent factors and (right) our CCR2 fibroblast latent factors.

**Figure S11 – Expression of the genes of interest in HF macrophages residing in spatial microniches (Accompanies Figure 5)**

Gene expression in HF macrophages residing in spatial microniches defined by proximity to activated or resting fibroblasts. * *p*<0.05, ** *p*<0.01, *** p<0.001, 2-tailed Student’s t-test

**Figure S12 – Expression of the genes and TFs of interest in sham vs MI cardiac macrophages (Accompanies Figure 6)**

Expression of B3GALNT1, TVP23B, TDRD6, MAZ, and MAFK in sham, MI mouse hearts by immunostaining, A-E: representative images), F: quantification of the latent factors. * *p*<0.05, ** *p*<0.01, *** p<0.001, **** *p*<0.0001, 2-tailed Student’s t-test.

**Figure S13 – Enumeration of leukocyte populations in mice after in vivo silencing of the selected genes and TFs (Accompanies Figure 6)**

Leukocyte populations in the blood (A), bone marrow (B), and spleen (C) of *siControl*, *siB3galnt1*, *siMafk*, *siMaz*, *siTdrd6*, and *siTvp23b* in mice two months after MI.

## Notes

### Competing Interest Statement

The authors have declared no competing interest.

