## Supplementary Figures for "Interpretable machine learning coupled to spatial transcriptomics unveils mechanisms of macrophage-driven fibroblast activation in ischemic cardiomyopathy"

Figure S1

**A**

| | |
|---|---|
| ischemic heart failure | n=4 |
| control | n=4 |

**B**

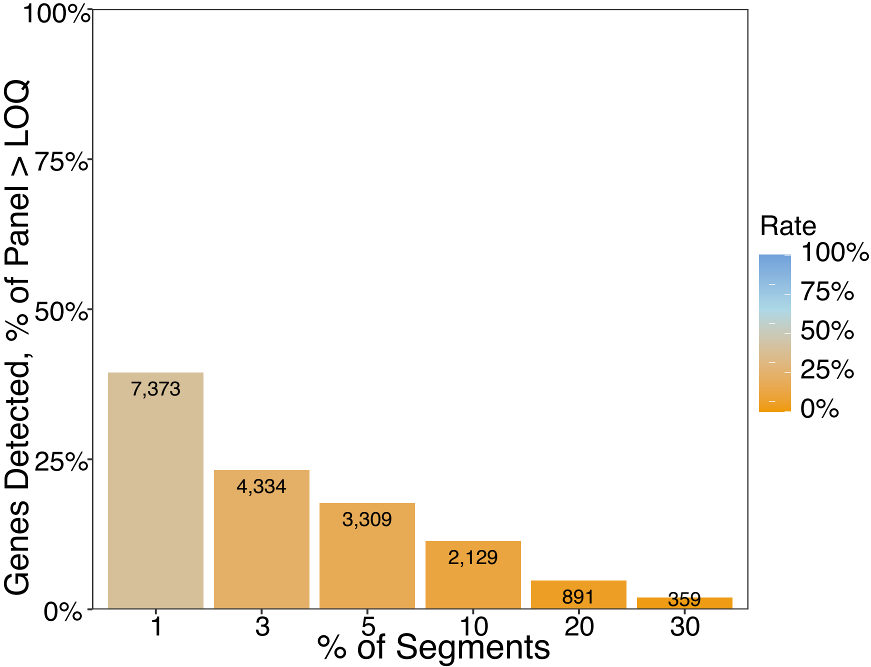

Figure S2

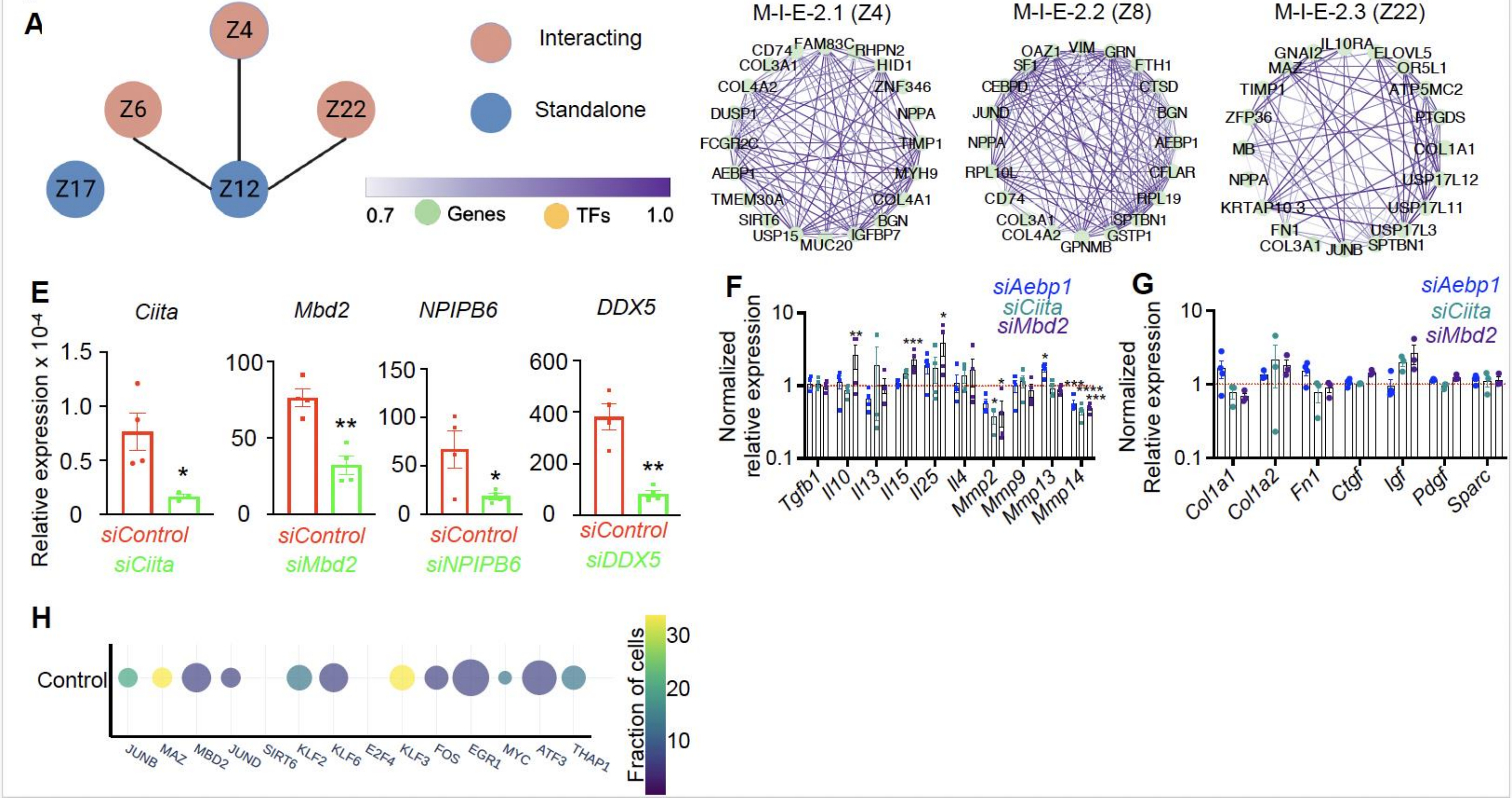
A
Z4
Z6
Z22
Z17
Z12
Interacting
Standalone
0.7
Genes
TFs
1.0
B: Cell Adhesion
M-I-E-2.1 (Z4)
CD74
FAM83C
RHPN2
COL3A1
HID1
COL4A2
ZNF346
DUSP1
NPPA
FCGR2C
TIMP1
AEBP1
MYH9
TMEM30A
COL4A1
SIRT6
BGN
USP15
MUC20
IGFBP7
C: Tissue repair
M-I-E-2.2 (Z8)
OAZ1
VIM
GRN
SF1
FTH1
CEBPD
CTSD
JUND
BGN
NPPA
AEBP1
RPL10L
CFLAR
CD74
RPL19
COL3A1
SPTBN1
COL4A2
GSTP1
GPNMB
D: Inflammatory response
M-I-E-2.3 (Z22)
GNAI2
IL10RA
ELOVL5
MAZ
OR5L1
TIMP1
ATP5MC2
ZFP36
PTGDS
MB
COL1A1
NPPA
USP17L12
KRTAP10.3
USP17L11
FN1
USP17L3
COL3A1
JUNB
SPTBN1
E
Relative expression x 10⁻⁴
Ciita
Mbd2
NPIPB6
DDX5
siControl
siCiita
siMbd2
siNPIPB6
siDDX5
F
Normalized relative expression
siAebp1
siCiita
siMbd2
Tgfb1
Il10
Il13
Il15
Il25
Il4
Mmp2
Mmp9
Mmp13
Mmp14
G
Normalized Relative expression
Col1a1
Col1a2
Fn1
Ctgf
Igf
Pdgf
Sparc
H
Control
JUNB
MAZ
MBD2
JUND
SIRT6
KLF2
KLF6
E2F4
KLF3
FOS
EGR1
MYC
ATF3
THAP1
Fraction of cells

Figure S3

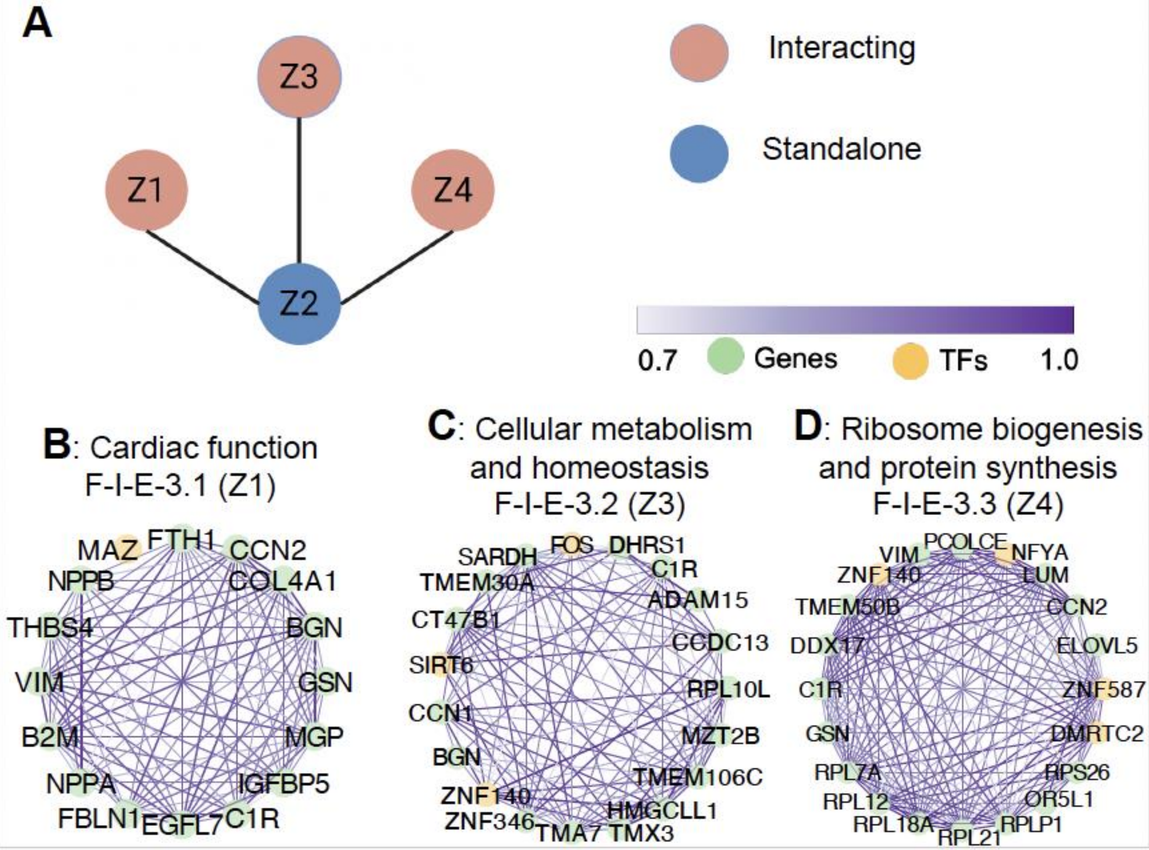

A
Z3
Z1
Z4
Z2
Interacting
Standalone
0.7
Genes
TFs
1.0
B: Cardiac function
F-I-E-3.1 (Z1)
MAZ
FTH1
CCN2
NPPB
COL4A1
THBS4
BGN
VIM
GSN
B2M
MGP
NPPA
IGFBP5
FBLN1
EGFL7
C1R
C: Cellular metabolism
and homeostasis
F-I-E-3.2 (Z3)
SARDH
FOS
DHRS1
C1R
TMEM30A
ADAM15
CT47B1
CCDC13
SIRT6
RPL10L
CCN1
MZT2B
BGN
TMEM106C
ZNF140
HMGCLL1
ZNF346
TMA7
TMX3
D: Ribosome biogenesis
and protein synthesis
F-I-E-3.3 (Z4)
VIM
PCOLCE
NFYA
ZNF140
LUM
TMEM50B
CCN2
DDX17
ELOVL5
C1R
ZNF587
GSN
DMRTC2
RPL7A
RPS26
RPL12
OR5L1
RPL18A
RPL21
RPLP1

Figure S4

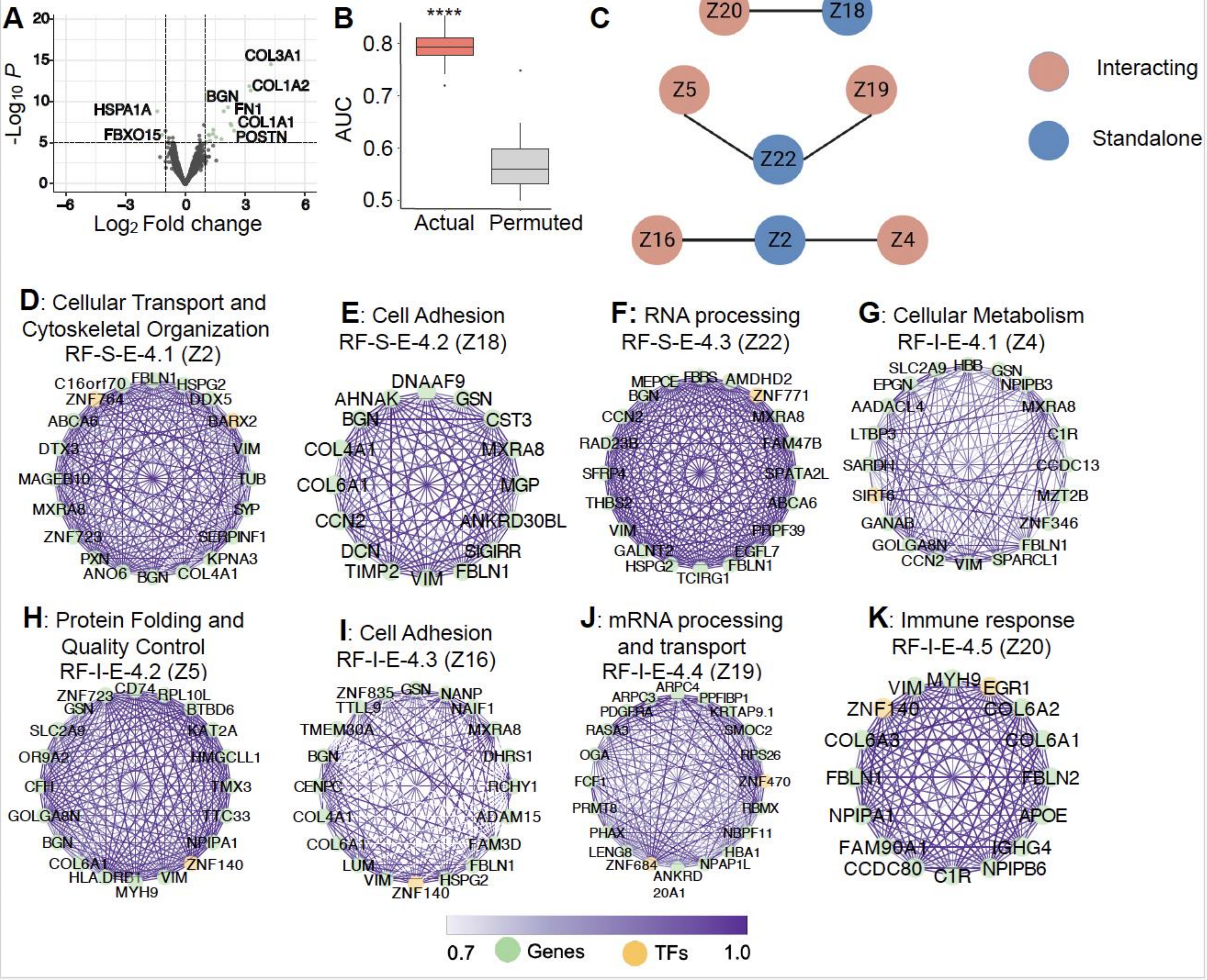

A
-Log10 P
Log2 Fold change
COL3A1
COL1A2
BGN
FN1
COL1A1
POSTN
HSPA1A
FBXO15
B
AUC
****
Actual
Permuted
C
Z20
Z18
Z5
Z19
Z22
Z16
Z2
Z4
Interacting
Standalone
D: Cellular Transport and Cytoskeletal Organization RF-S-E-4.1 (Z2)
C16orf70 FBLN1 HSPG2 ZNF764 DDX5 ABCA6 BARX2 DTX3 VIM MAGEB10 TUB MXRA8 SYP ZNF723 SERPINF1 PXN KPNA3 ANO6 BGN COL4A1
E: Cell Adhesion RF-S-E-4.2 (Z18)
DNAAF9 AHNAK GSN BGN CST3 COL4A1 MXRA8 COL6A1 MGP CCN2 ANKRD30BL DCN SIGIRR TIMP2 VIM FBLN1
F: RNA processing RF-S-E-4.3 (Z22)
MEPCE FBRS AMDHD2 BGN ZNF771 CCN2 MXRA8 RAD23B FAM47B SFRP4 SPATA2L THBS2 ABCA6 VIM PRPF39 GALNT2 EGFL7 HSPG2 FBLN1 TCIRG1
G: Cellular Metabolism RF-I-E-4.1 (Z4)
SLC2A9 HBB GSN EPGN NPIPB3 AADACL4 MXRA8 LTBP3 C1R SARDH CCDC13 SIRT6 MZT2B GANAB ZNF346 GOLGA8N FBLN1 CCN2 VIM SPARCL1
H: Protein Folding and Quality Control RF-I-E-4.2 (Z5)
ZNF723 CD74 RPL10L GSN BTBD6 SLC2A9 KAT2A OR9A2 HMGCLL1 CFH TMX3 GOLGA8N TTC33 BGN NPIPA1 COL6A1 ZNF140 HLA.DRB1 VIM MYH9
I: Cell Adhesion RF-I-E-4.3 (Z16)
ZNF835 GSN NANP TTLL9 NAIF1 TMEM30A MXRA8 BGN DHRS1 CENPC RCHY1 COL4A1 ADAM15 COL6A1 FAM3D LUM FBLN1 VIM HSPG2 ZNF140
J: mRNA processing and transport RF-I-E-4.4 (Z19)
ARPC4 ARPC3 PPFIBP1 PDGFRA KRTAP9.1 RASA3 SMOC2 OGA RPS26 FCF1 ZNF470 PRMT8 RBMX PHAX NBPF11 LENG8 HBA1 ZNF684 NPAP1L ANKRD 20A1
K: Immune response RF-I-E-4.5 (Z20)
VIM MYH9 EGR1 ZNF140 COL6A2 COL6A3 COL6A1 FBLN1 FBLN2 NPIPA1 APOE FAM90A1 IGHG4 CCDC80 C1R NPIPB6
0.7
Genes
TFs
1.0

Figure S5 - Activated Fibroblasts

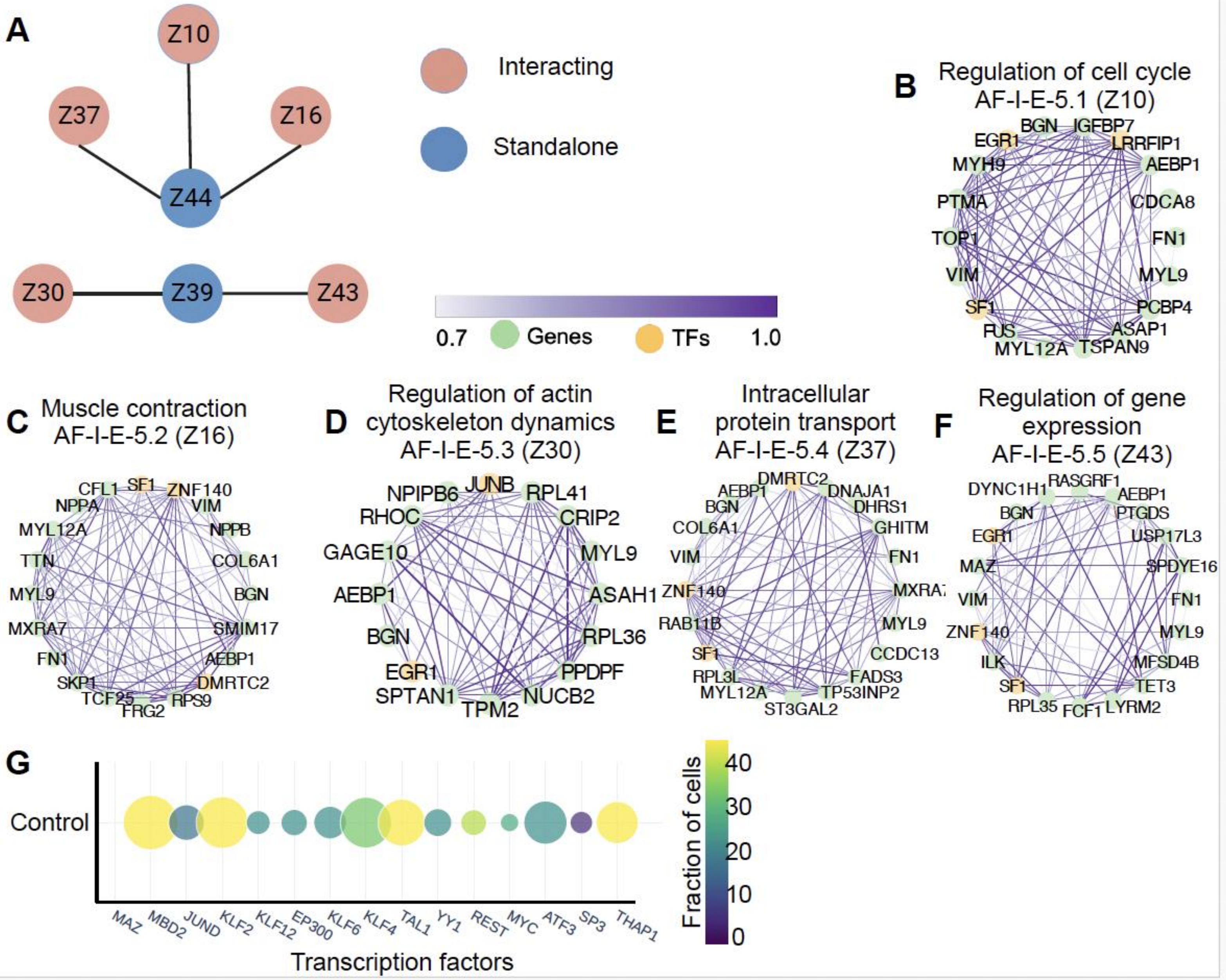

Figure S6 - Macrophages near activated fibroblasts

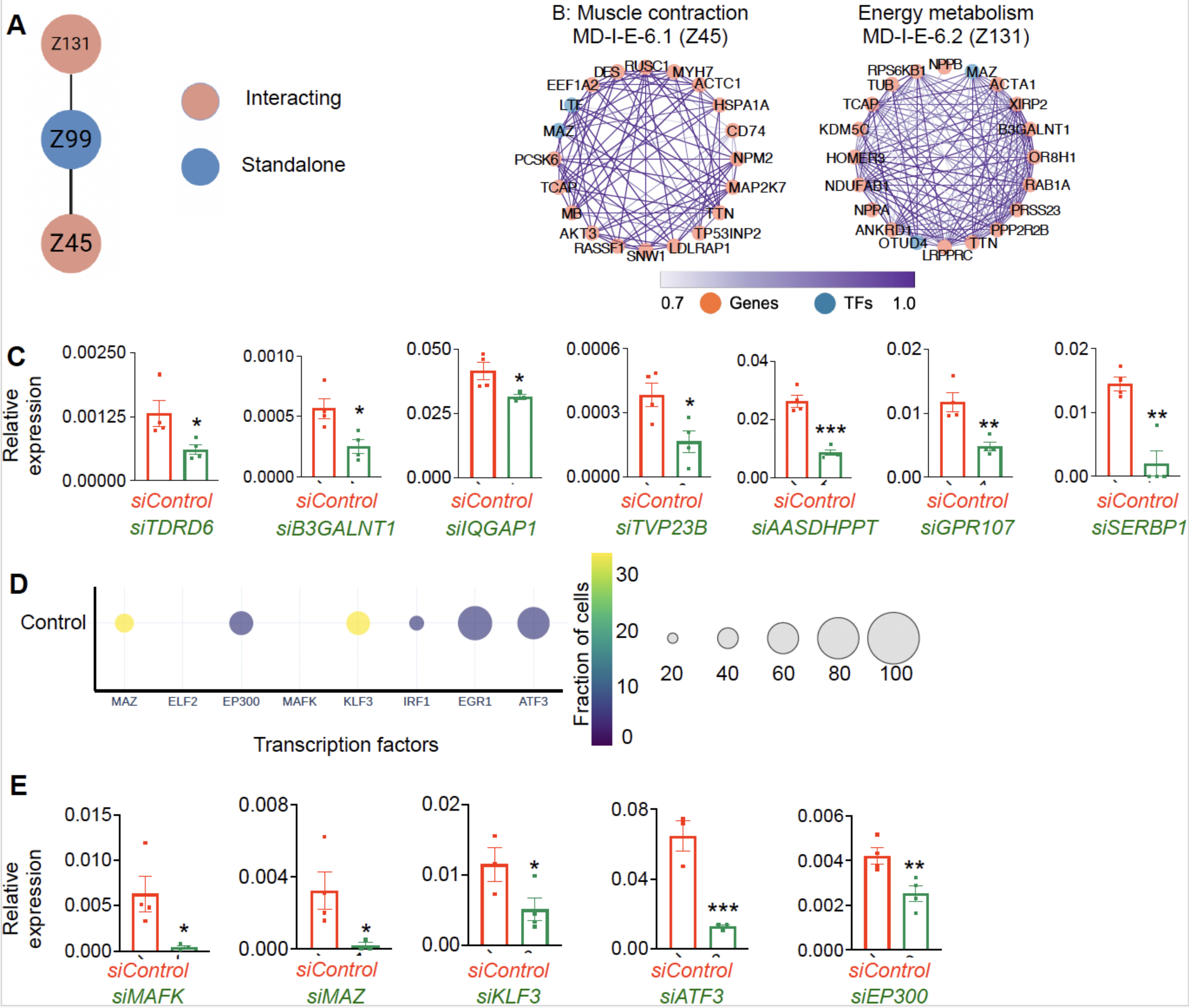

Figure S7

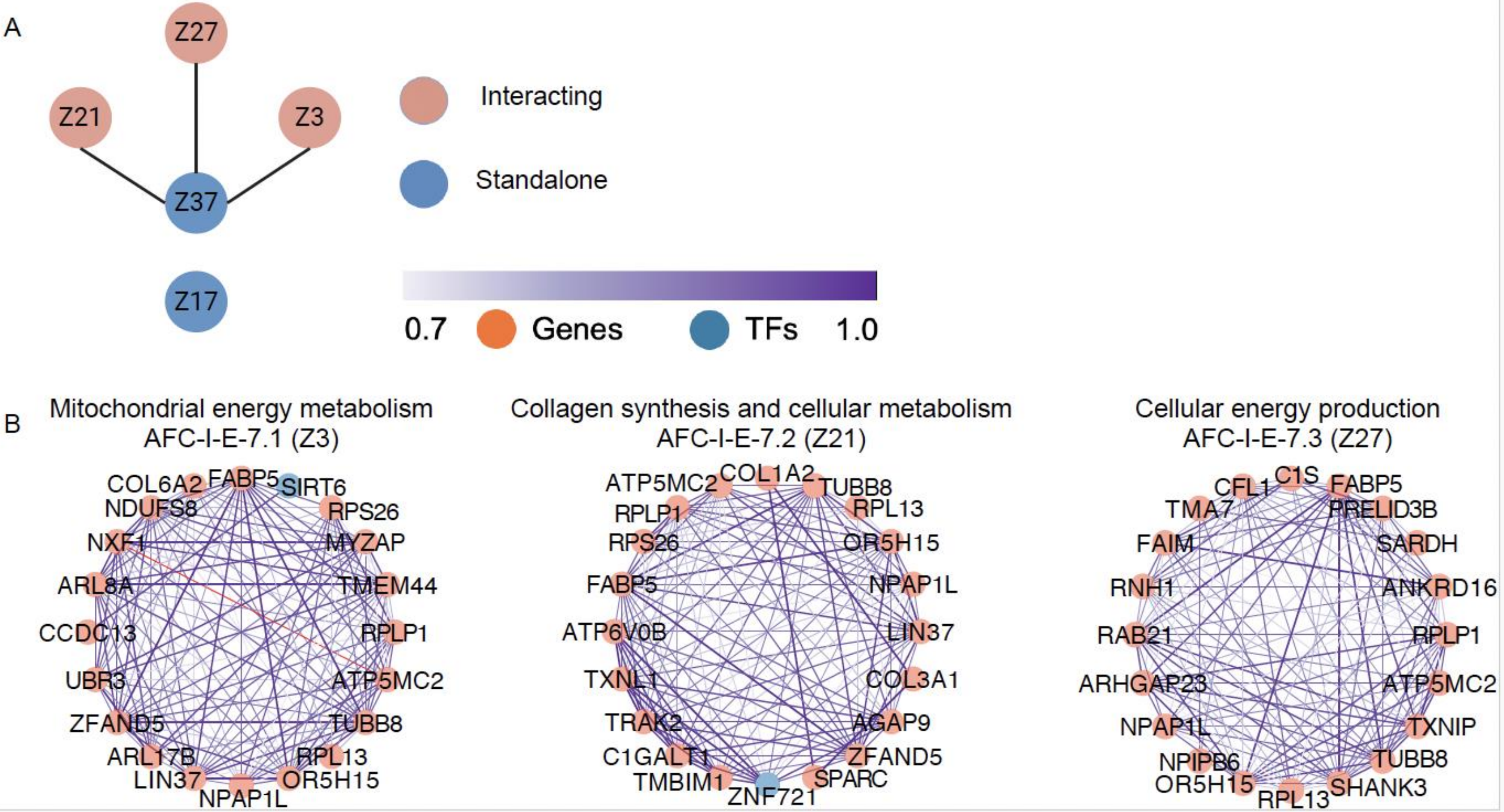
A
Z27
Z21
Z3
Z37
Z17
Interacting
Standalone
0.7
Genes
TFs
1.0
B
Mitochondrial energy metabolism
AFC-I-E-7.1 (Z3)
COL6A2
FABP5
SIRT6
NDUFS8
RPS26
NXF1
MYZAP
ARL8A
TMEM44
CCDC13
RPLP1
UBR3
ATP5MC2
ZFAND5
TUBB8
ARL17B
RPL13
LIN37
OR5H15
NPAP1L
Collagen synthesis and cellular metabolism
AFC-I-E-7.2 (Z21)
ATP5MC2
COL1A2
TUBB8
RPLP1
RPL13
RPS26
OR5H15
FABP5
NPAP1L
ATP6V0B
LIN37
TXNL1
COL3A1
TRAK2
AGAP9
C1GALT1
ZFAND5
TMBIM1
SPARC
ZNF721
Cellular energy production
AFC-I-E-7.3 (Z27)
CFL1
C1S
FABP5
TMA7
PRELID3B
FAIM
SARDH
RNH1
ANKRD16
RAB21
RPLP1
ARHGAP23
ATP5MC2
NPAP1L
TXNIP
NPIPB6
TUBB8
OR5H15
SHANK3
RPL13

Figure S8

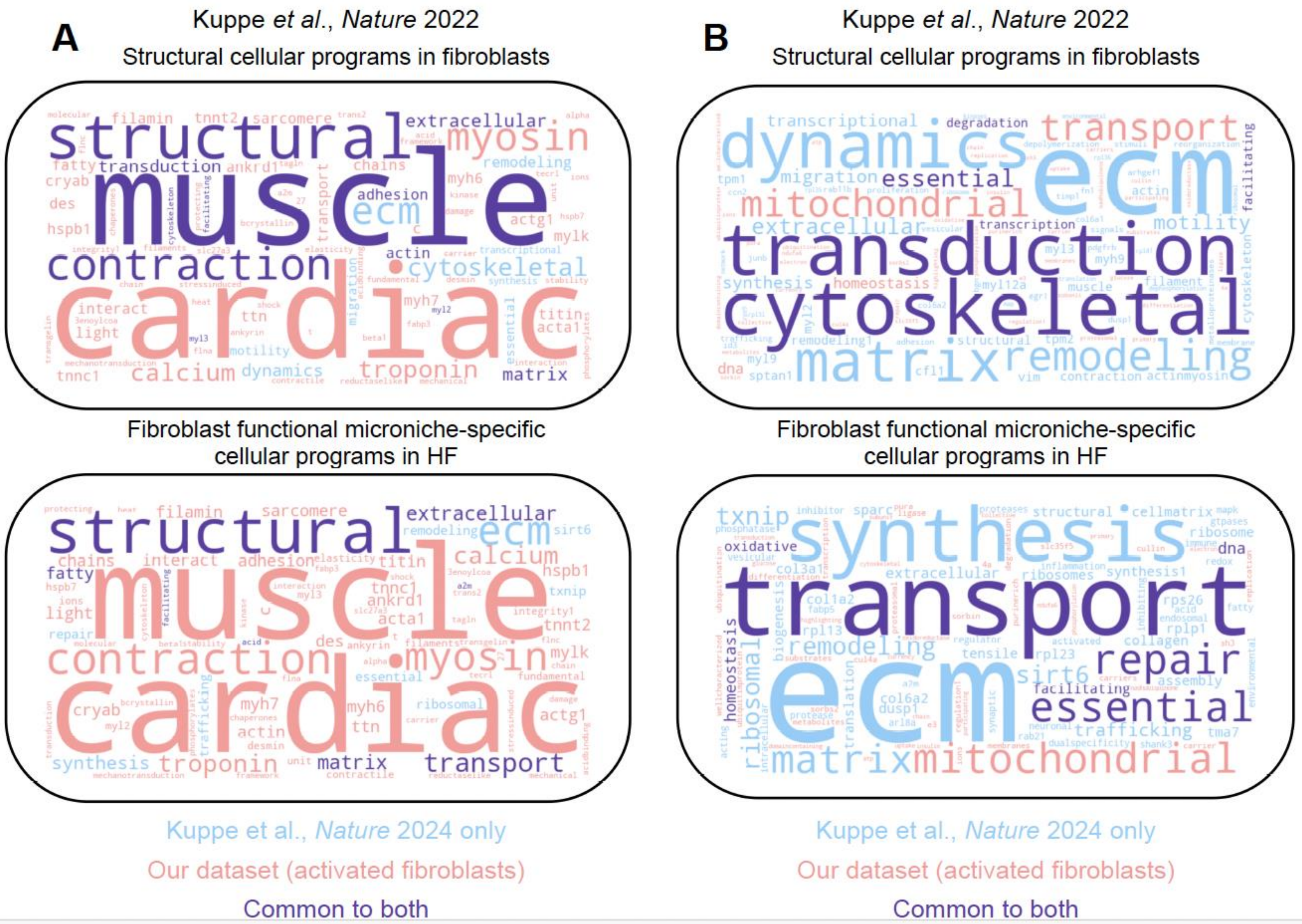
A
Kuppe et al., Nature 2022
Structural cellular programs in fibroblasts
structural
muscle
cardiac
contraction
myosin
extracellular
cytoskeletal
ecm
troponin
calcium
Fibroblast functional microniche-specific cellular programs in HF
structural
muscle
cardiac
contraction
myosin
extracellular
ecm
calcium
troponin
transport
matrix
B
Kuppe et al., Nature 2022
Structural cellular programs in fibroblasts
dynamics
ecm
transport
mitochondrial
transduction
cytoskeletal
matrix
remodeling
Fibroblast functional microniche-specific cellular programs in HF
synthesis
transport
ecm
repair
essential
mitochondrial
matrix
Kuppe et al., Nature 2024 only
Our dataset (activated fibroblasts)
Common to both

Figure S9

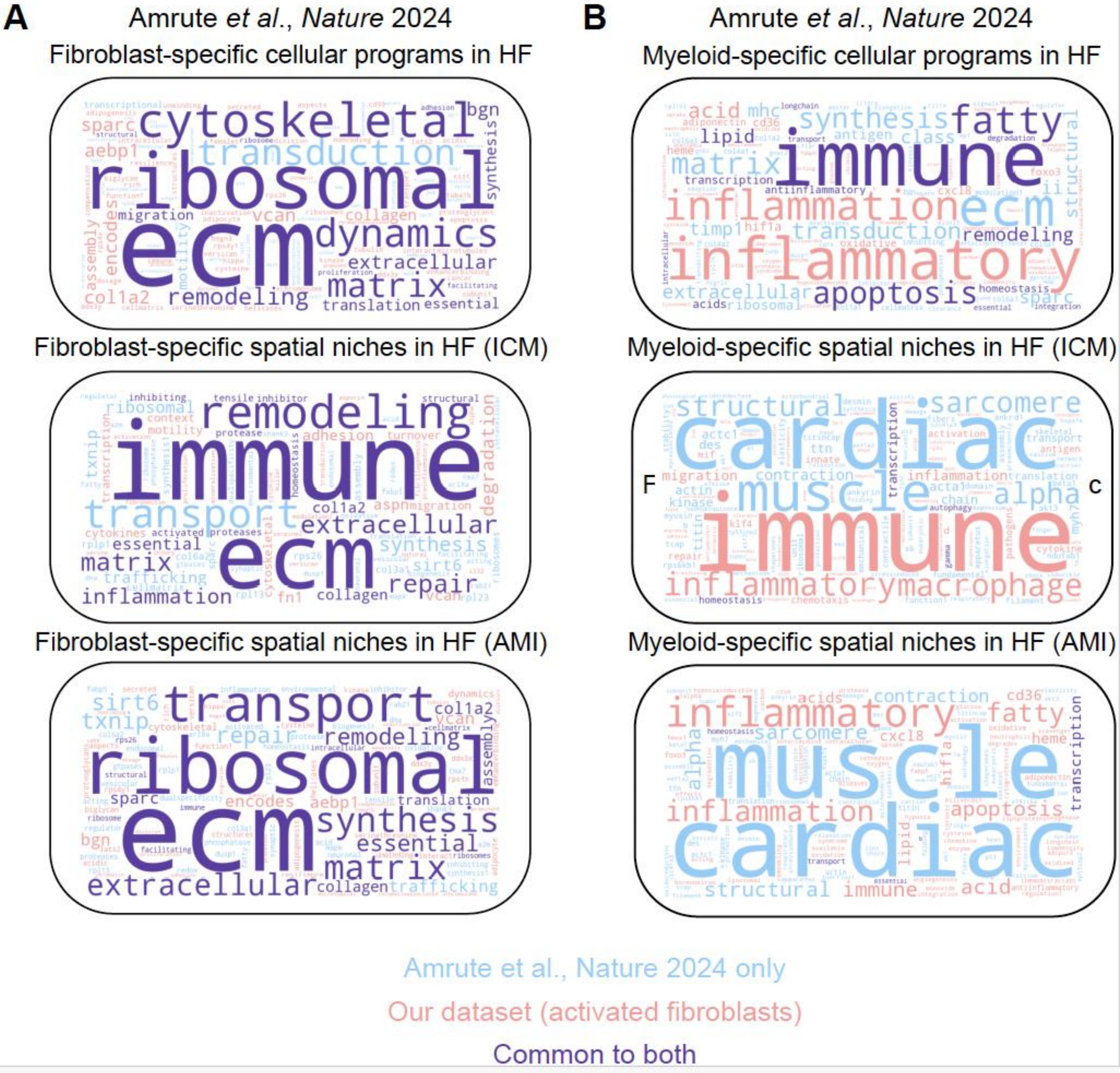

A
Amrute et al., Nature 2024
Fibroblast-specific cellular programs in HF
Fibroblast-specific spatial niches in HF (ICM)
Fibroblast-specific spatial niches in HF (AMI)
B
Amrute et al., Nature 2024
Myeloid-specific cellular programs in HF
Myeloid-specific spatial niches in HF (ICM)
Myeloid-specific spatial niches in HF (AMI)
Amrute et al., Nature 2024 only
Our dataset (activated fibroblasts)
Common to both

Figure S10

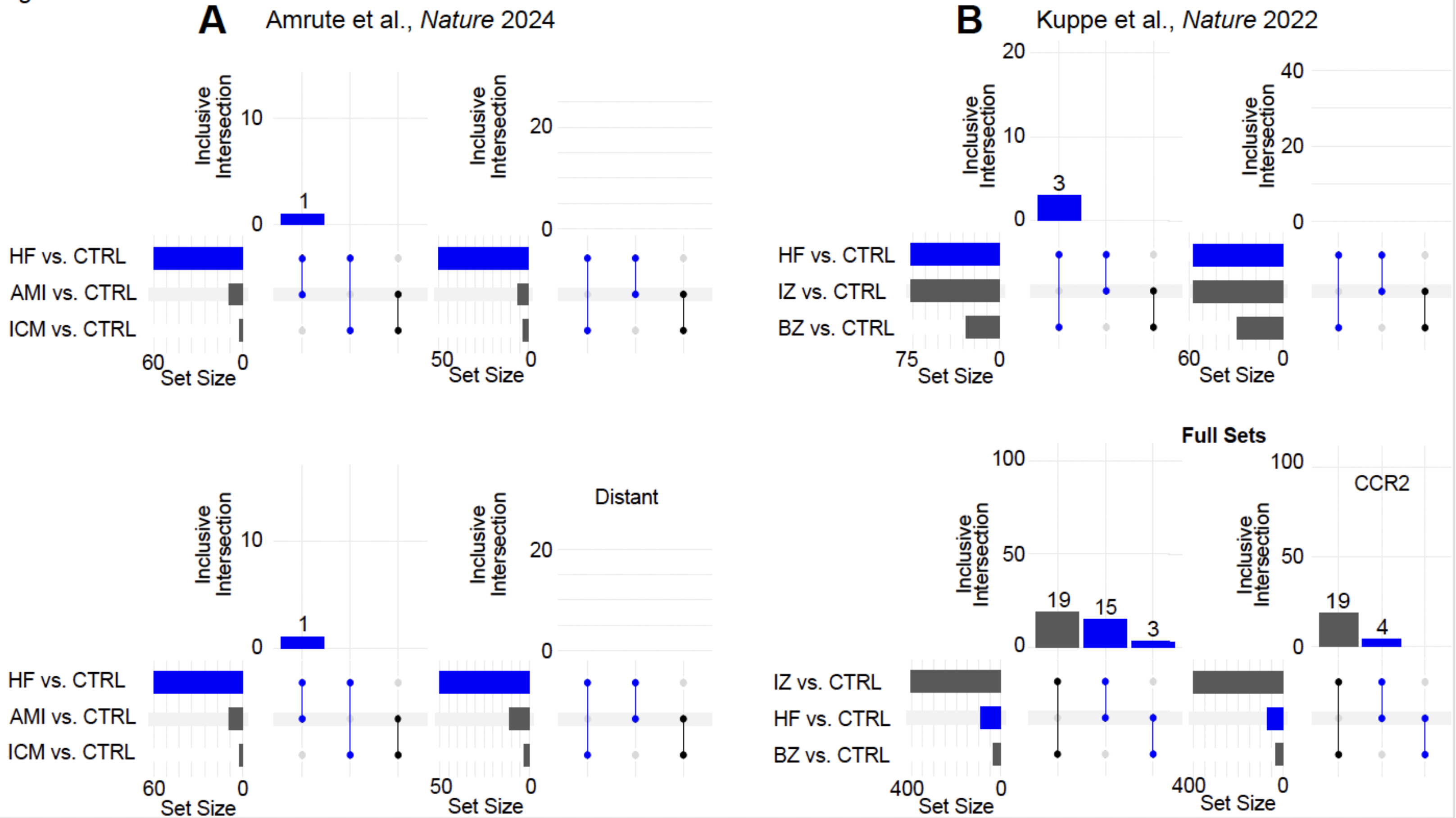

A
Amrute et al., Nature 2024
B
Kuppe et al., Nature 2022
Inclusive Intersection
HF vs. CTRL
AMI vs. CTRL
ICM vs. CTRL
IZ vs. CTRL
BZ vs. CTRL
Set Size
Distant
Full Sets
CCR2

Figure S11

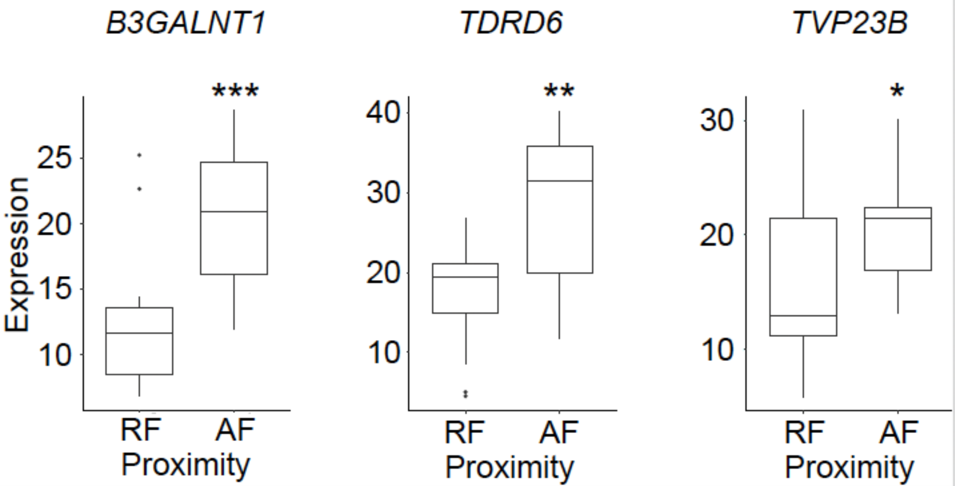

B3GALNT1
TDRD6
TVP23B
***
**
*
Expression
25
20
15
10
40
30
20
10
30
20
10
RF
AF
Proximity
RF
AF
Proximity
RF
AF
Proximity

Figure S12

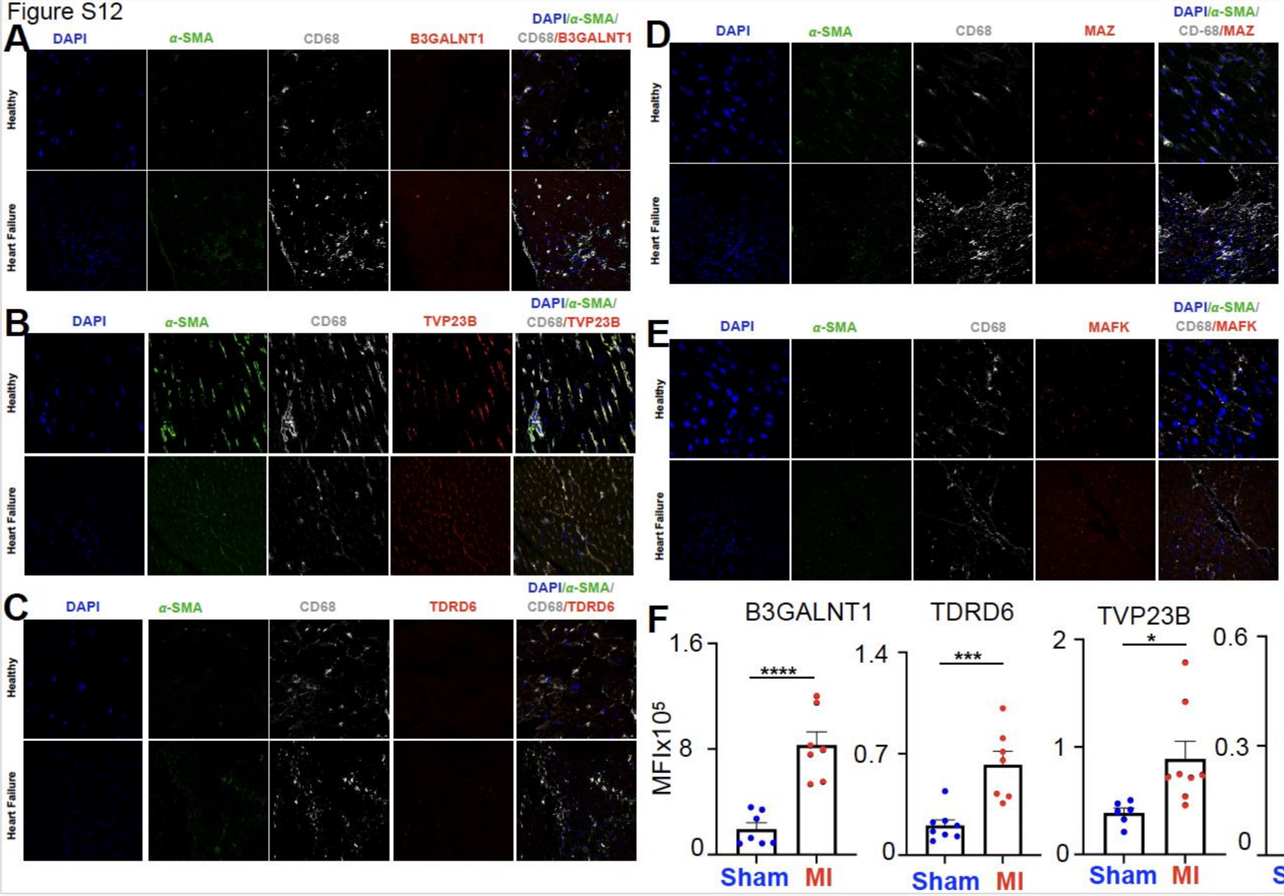
A
DAPI
α-SMA
CD68
B3GALNT1
DAPI/α-SMA/CD68/B3GALNT1
Healthy
Heart Failure
B
DAPI
α-SMA
CD68
TVP23B
DAPI/α-SMA/CD68/TVP23B
Healthy
Heart Failure
C
DAPI
α-SMA
CD68
TDRD6
DAPI/α-SMA/CD68/TDRD6
Healthy
Heart Failure
D
DAPI
α-SMA
CD68
MAZ
DAPI/α-SMA/CD-68/MAZ
Healthy
Heart Failure
E
DAPI
α-SMA
CD68
MAFK
DAPI/α-SMA/CD68/MAFK
Healthy
Heart Failure
F
B3GALNT1
TDRD6
TVP23B
MAZ
MAFK
MFIx10^5
****
***
*
ns
*
1.6
8
0
1.4
0.7
0
2
1
0
0.6
0.3
0
1.6
0.8
0
Sham
MI

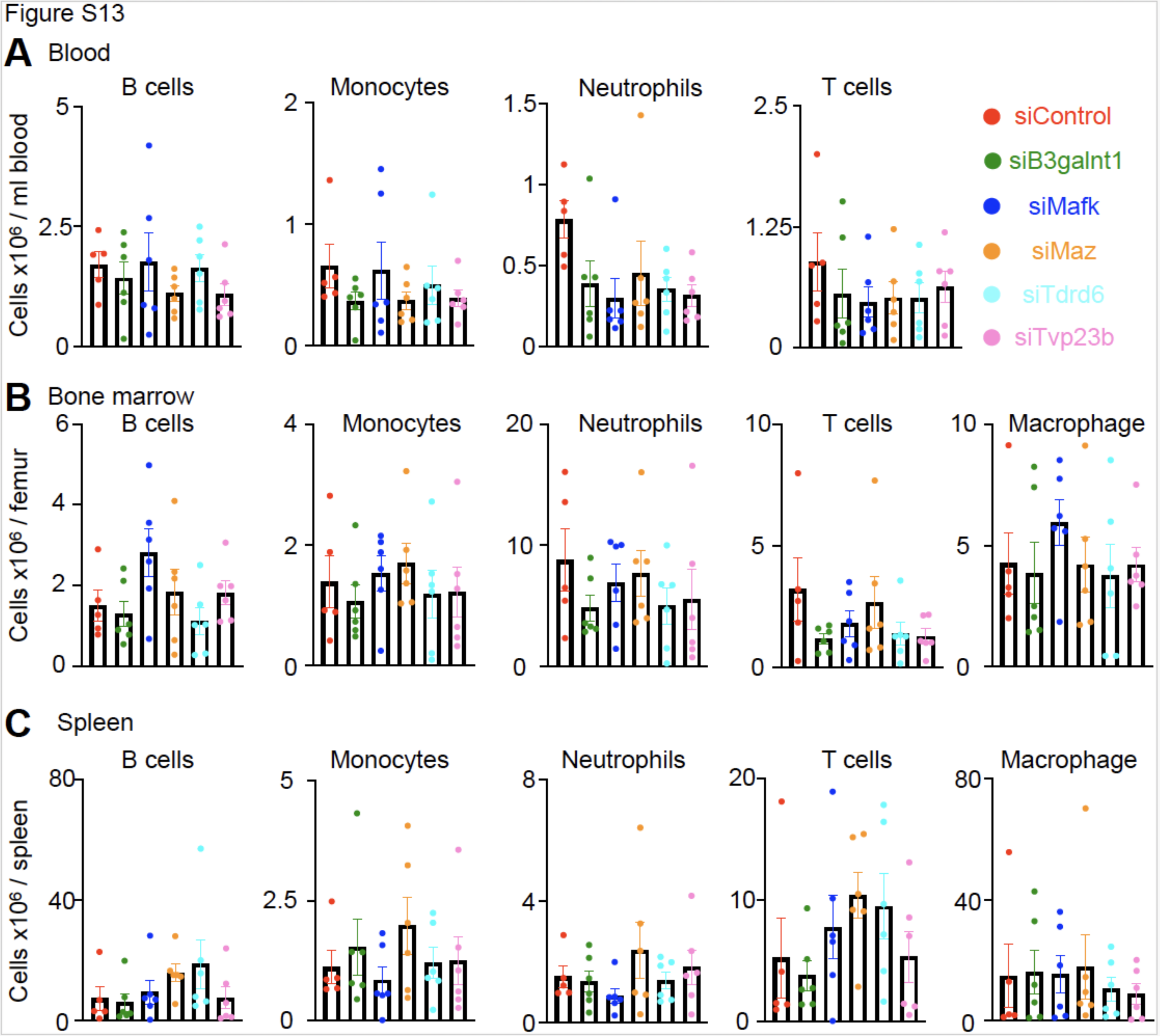
Figure S13
A Blood
B cells
Monocytes
Neutrophils
T cells
Cells x10⁶ / ml blood
siControl
siB3galnt1
siMafk
siMaz
siTdrd6
siTvp23b
B Bone marrow
B cells
Monocytes
Neutrophils
T cells
Macrophage
Cells x10⁶ / femur
C Spleen
B cells
Monocytes
Neutrophils
T cells
Macrophage
Cells x10⁶ / spleen
